# COVID-19 epidemic in Malaysia: Impact of lockdown on infection dynamics

**DOI:** 10.1101/2020.04.08.20057463

**Authors:** Naomie Salim, Weng Howe Chan, Shuhaimi Mansor, Nor Erne Nazira Bazin, Safiya Amaran, Ahmad Athif Mohd Faudzi, Anazida Zainal, Sharin Hazlin Huspi, Eric Khoo Jiun Hooi, Shaekh Mohammad Shithil

## Abstract

COVID-19 epidemic in Malaysia started as a small wave of 22 cases in January 2020 through imported cases. It was followed by a bigger wave mainly from local transmissions resulting in 651 cases. The following wave saw unexpectedly three digit number of daily cases following a mass gathering urged the government to choose a more stringent measure. A limited lock-down approach called Movement Control Order (MCO) was immediately initiated to the whole country as a way to suppress the epidemic trajectory. The lock-down causes a major socio-economic disruption thus the ability to forecast the infection dynamic is urgently required to assist the government on timely decisions. Limited testing capacity and limited epidemiological data complicate the understanding of the future infection dynamic of the COVID-19 epidemic. Three different epidemic forecasting models was used to generate forecasts of COVID-19 cases in Malaysia using daily reported cumulative case data up until 1st April 2020 from the Malaysia Ministry of Health. The forecasts were generated using a Curve Fitting Model with Probability Density Function and Skewness Effect, the SIR Model, and a System Dynamic Model. Method one based on curve fitting with probability density function estimated that the peak will be on 19th April 2020 with an estimation of 5,637 infected persons. Method two based on SIR Model estimated that the peak will be on 20th - 31st May 2020 if Movement Contro (MCO) is in place with an estimation of 630,000 to 800,000 infected persons. Method three based on System Dynamic Model estimated that the peak will be on 17th May 2020 with an estimation of 22,421 infected persons. Forecasts from each of model suggested the epidemic may peak between middle of April to end of May 2020.

## 1. Introduction

A novel coronavirus infectious disease (COVID-19) which is caused by SARS-CoV-2 has been announced by the World Health Organization as a fatal global pandemic [1]. The epidemic of COVID- 19 started explosively in Wuhan and spread throughout China [2]. Mediated via a massive aviation industry, it turned into a pandemic in just two months [3]. As of April 4, 2020, the number of cases climbed above 1 million with a death toll of over 50 000 worldwide [1]. The global impact and the public health threat of COVID-19 is the most serious seen in a respiratory virus since the 1918 influenza pandemic [4]. Both COVID-19 and the 1918 influenza pandemic are associated with respiratory spread, a significant percentage of infected people with asymptomatic cases transmitting infection to others, and a high fatality rate [5-6].

Globally, the epidemic curve in each country varies from exponential, uncontrolled outbreak (Italy) to slow rising, adequately controlled (Singapore) [1]. Malaysia somehow lies in the middle [1]. As of April 5, 2020, there are 3,662 cases and 61 deaths in Malaysia [7]. Malaysian government are taking prompt public health actions to prevent an exponential rise of cases by continuously screen and test high risk individuals, isolate patients and trace and quarantine the contacts to prevent secondary spread [8]. These actions seemed to be adequate until a large cluster of cases occurred following a large Tabligh gathering involving more than 10,000 members in late February [9]. The event has changed the direction of the epidemic curve in Malaysia [10].

The dire urgency in controlling the outbreak to prevent the collapse of healthcare system has forced the government to impose a more stringent action [11]. Malaysian Prime Minister announced a limited lock-down called the Movement Control Order (MCO) on 16th March 2020. The first MCO (MCO1) started on 18th March until 31st March 20207. It was then continued for another 2 weeks (MCO2) until 14th April 2020 [12].

During MCO, all universities, schools, religious places and non-essential sectors are closed. Inter- state travel are not allowed unless for valid reasons. Only the head of family is allowed to buy groceries within 10 km radius. Both police and army works together in coordinating and monitoring peoples’ movements.

This stringent action is not without a cost to society. It has a major social and economic disruptions [11-12]. The uncertainties related to the outbreak also creates anxiety. Although the decision to implement MCO was acceptable to many in view of outbreak casualties, but the question is how many weeks is needed? A study showed if we lift the suppressive measure too early, the massive outbreak may recur. On the contrary, if the measure was in place for too long, the social, economic, and psychological effect will be massive [12].

Simple counts of the number of confirmed cases can be misleading indicators of the epidemic’s trajectory if these counts are limited by problems in access to care or bottlenecks in laboratory testing, or if only patients with symptoms are tested [2]. This is where a prediction modelling may assist the authority in making decisions. Model-based predictions can help policy makers make the right decisions in a timely way, even with the uncertainties about COVID-19.

Therefore, in this work, we try to predict the projection of COVID-19 outbreak cases in Malaysia using three mathematical models; Curve Fitting Model with Probability Density Function and Skewness Effect, SIR model, System Dynamic model. We used a combination of actual daily data and analysis of patterns and trends from previous cases in other countries to predict the projection of upcoming cases for Malaysia. The projection serves to support the needs for lockdown period and activity to mitigate the spread of coronavirus cases. Accurate prediction is very crucial to support right decision for upcoming lockdown period and activity. For instance, extended period for MCO can be decided based on increasing and descending trends of COVID-19 cases.

This paper is organized in five sections. Section 2 reviews some prediction modelling concepts and their related works. Section 3 describes the three prediction methods (Curve Fitting Model, SIR Model and System Dynamics) used in this study and their experimental assumptions made. Finally, Section 4 presents results obtained by each of these prediction models and Section 5 concludes the paper.

## 2. Related Work

Many models have been used to predict the outbreak pattern of COVID-19 epidemic. Several models used the normal distribution as a model of the COVID-19 epidemic and to forecast peak hospital load [13]. A curve-fitting tool to fit a nonlinear mixed effects model was developed based on available data. For instance, the cumulative rate is assumed to follow a parameterized Gaussian error function where the function is the Gaussian error function. Parameters such as death rate, the time since death rate exceeded a certain number was used as a location-specific inflection point and location-specific growth parameter, have been used. Other sigmoidal functional forms were also considered. Data was also fit to the log of the death rate in the available data, using an optimization framework. The logistic distribution is based on a continuous probability distribution which resembles the normal distribution in shape but has heavier tails. A generalized logistic growth model was used by [14], together with the Richards growth model and a sub-epidemic wave model to generate COVID-19 10-day forecasts for Guangdong and Zhejiang. The generalized logistic growth model and the Richards model extend the simple logistic growth model with an additional scaling parameter. The sub-epidemic model accommodates complex epidemic trajectories by assembling the contribution of inferred overlapping sub-epidemics. The model was fit to the “incidence” curve and the best-fit solution for each model was estimated using nonlinear least squares fitting so that model parameters minimizes the sum of squared errors between the model and the data. A parametric bootstrap approach was used to generate uncertainty bounds around the best-fit solution assuming a Poisson error structure.

The SIR model is one of the simplest models to predict properties of how a disease spreads, for example total number of infected or the duration of an epidemic. The population is first divided into compartments, with the assumption that every individual in the same compartment has the same characteristics [15]. The model consists of three compartments: S for the number of susceptible, I for the number of infectious, and R for the number of recovered or deceased (or immune) individuals. This model is reasonably predictive for infectious diseases which are transmitted from human to human. During an epidemic, the number of susceptible individual falls rapidly as more of them are infected and thus enter the infectious and recovered compartments. Each member of the population typically progresses from susceptible to infectious to recovered. JP Morgan used a model based on SIR to estimate the COVID-19 epidemic curve in Malaysia [16]. The estimation is based on the potential size of the group that initially interacts with the infected group (i.e., group needs to get the virus test) to be around ∼0.2% of the total population based on the total size of the test group in China’s Hubei and South Korea, which is about 0.1 and 0.7% of the total population, respectively. The secondary infection rate (R0) was adopted based on infection parameters used in China (2) and South Korea (1.9) where a country would face a doubling of infection process in every 5-7 days in the early acceleration stage. Due to several containment measures and lower population density of Malaysia (96 people per sq. km of land area vs. Japan/ Korea: 347/ 212 according to World Bank), they set 1.7 as the initial setting of R0. Imperial College COVID-19 Response Team modified an individual-based simulation model for pandemic influenza planning by [17] to explore scenarios for COVID-19 in Great Britain. For their model, they assumed an incubation period of 5.1 days, infectiousness to occur from 12 hours prior to the onset of symptoms for those that are symptomatic and from 4.6 days after infection in those that are asymptomatic with an infectiousness profile over time that results in a 6.5-day mean generation time. They used R0=2.4 based on fits to the early growth-rate of the epidemic in Wuhan and symptomatic individuals as 50% more infectious than asymptomatic individuals. On recovery from infection, individuals are assumed to be immune to re-infection in the short term. In addition, Pueyo [18] uses the epidemic calculator provided by Goh [19] to predict the effect of control measures on spread of infection in the United States and how it will affect their healthcare services. Chen et al. proposed a time-delay dynamic system based on five compartments: external suspected people, infected people, confirmed people, isolated people, and cured people [20]. They also added external sources such as, spread rate, latent period, delay period, exposed people, and cured rate to describe the trend of COVID-19 outbreak.

However, curve fitting models mentioned above expects data from a small portion of the behavior to predict the peak. If variations occur in the data, such as dramatic shifts in test coverage, the forecast might not be accurate. To solve this problem, other methods such as machine learning can be used to analyze information from a multitude of sources and track over a hundred infectious diseases (Forbes, 2020). For instance, in December 2019, Blue Dot predicted the COVID19 outbreak using machine learning and sent out a warning to its customers to avoid Wuhan, ahead of both the US Centers for Disease Control and Prevention (CDC) and the World Health Organization (WHO). Blue Dot also predicted where other Asian city outbreaks could be by analyzing traveler itineraries and flight paths. However insufficient amount of available data when an epidemic just started is a big challenge in machine learning. In the past, three popular methods have been proposed, they include 1) augmenting the existing little data, 2) using a panel selection to pick the best forecasting model from several models, and 3) fine-tuning the parameters of an individual forecasting model for the highest possible accuracy. Fong et al. proposed a methodology based on data augmentation to the existing little data, panel selection to pick the best forecasting model from several models and fine-tuning the parameters of an individual forecasting model for the highest possible accuracy [21]. They constructed a polynomial neural network with corrective feedback model to forecast the COVID-19 outbreak with low prediction error, which is useful for predicting disease outbreak when the samples are small.

In terms of data, earliest data on COVID-19 are from China, with case fatalities as high as 1% among the infected [22]. The global mortality ratio averaged at around 4.4% whilst Malaysia’s reported mortality ratio is 0.77% [16]. There are several data repository for COVID19. Examples are data available from Worldometer [23] and the 2019 Novel Coronavirus Visual Dashboard [24] operated by the Johns Hopkins University Center for Systems Science and Engineering (JHU CSSE), which is also supported by ESRI Living Atlas Team and the Johns Hopkins University Applied Physics Lab. For Malaysia, data has been published by the STAR [25] and Malaysiakini [26].

For Malaysia, JP Morgan forecasted a mid-April infection peak of 6,300 for Malaysia [16]. The lower peak could be due to Malaysia’s ‘test per million capita’ of 482, which is four to 81 times higher than its neighboring ASEAN countries (6 to 109 tests per million population) and higher than several EU countries. Malaysia has made efforts to control the infection curve with school closures, bans on social gatherings, both inbound/ outbound travel restrictions and enforced the Movement Control Order effective from 18th March 2020 It is important that the government take actions to control the peak to ensure that the health facilities can cope with the need for hospitalization. The proportion of infected cases that required hospitalization and ICU admission were 20.7%-31.4% and 4.9-11.5%, respectively [16]. Malaysia’s current critical care beds are estimated at 1,060. The impact on economy can also be estimated, as has been done by Pearce [27].

## 3 Datasets, Methods and Results

### 3.1 Datasets

In order for the statistical and machine learning algorithms to learn and predict the trend and growth of the disease, several online news and related websites (such as the Malaysia’s official health ministry websites) was crawled and fed into a database. Covid-19 data on the number of susceptible, infectious, recovered, and deceased patients for world countries are available from Worldometer [23] and the 2019 Novel Coronavirus Visual Dashboard [24]. For Malaysia, daily data has been published by the STAR [25], Malaysiakini [26] and also by the Ministry of Health Malaysia [8-10].

For the Malaysian COVID-19 dataset, data on medical capacity (e.g.: number of beds in each state) and events that could affect the spread of the disease (example: Tabligh Assembly at Sri Petaling Mosque) were also collected to see if this data could help in making a strategy to flatten the curve of infected cases. The world data on the dates of restriction and quarantine declared by each affected country were also collected in order to gauge and infer how the infection would pattern of COVID-19 cases in Malaysia looks like, if similar controlled measures are implemented.

### 3.2. Method 1: Curve Fitting with Probability Density Function and Skewness Effect Modelling

In this model, we used a statistical method based on normal distribution function based on probability density function incorporating a skewness effect to estimate the pattern and peak of COVID-19 spread in Malaysia. We divided the model into two phases as will be described in the following paragraphs. Phase 1 is modeled based on curve fitting to project the number of cases by day up to 15 April, 2020. Phase 2 uses probability density function to estimate projection for recovery period to flatten the curve.

#### 3.1.1. Phase 1: Projection method to estimate number of cases by 15th April 2020

Suspected Coronavirus cases in Malaysia began on the 22nd January. On 25th January four new cases were confirmed. The first MCO was implemented on the 18th March until 31st March. Due to high rate of growth cases within a week, instruction for another MCO was continued from 1st April to 15th April.

The first initiative is to study the current trends in order to predict the rate of growth and number of cases on the 15^th^ of April. Plotting of the actual cases of COVID-19 daily from 22^nd^ January until 1nd April is shown in Figure 1. The plot shows the rate of growth of total cases and recovered cases. To understand the overall scenario of the trends, the active cases (i.e., total cases minus recovered cases), new cases, recovered cases and death cases are plotted together. The increase rate of total cases is around 125 cases per day.

**Figure 1.**
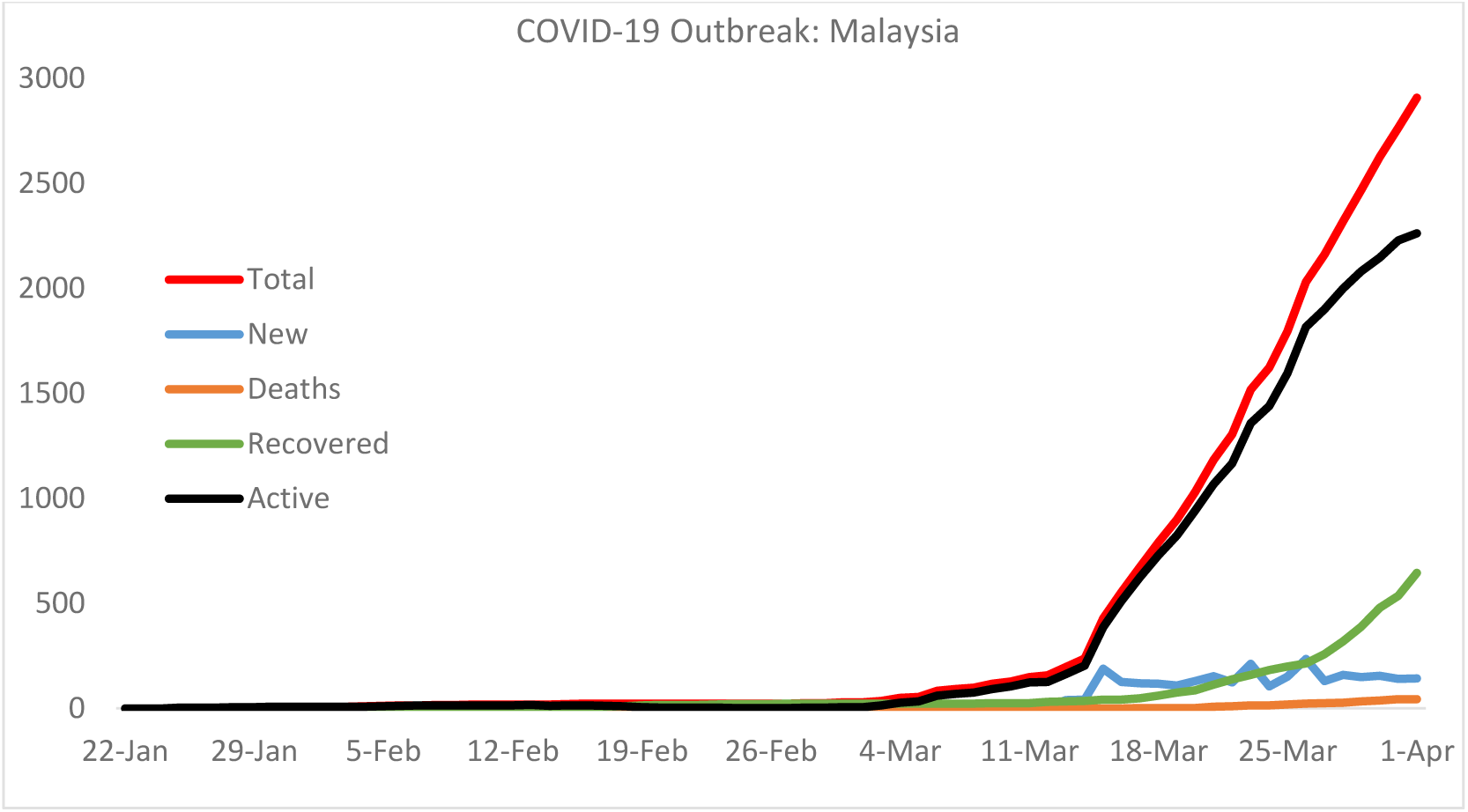
Progression of COVID-19 cases in Malaysia

The projection of total cases can be estimated from the trend-line equation generated through curve-fitting of the actual data using 5th order polynomial equation. The estimated trend line equation is given by the following equation.

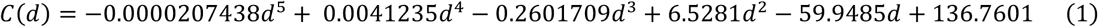

where *C* and *d* represents total cases and days respectively. The number of total cases by 15^th^ April 2020 is calculated by substituting number of days in Eq.1. The dotted line in Figure 2 shows the projection of total cases and it is estimated 5,637 of total cases in 85 days (i.e., number of days from 22^nd^ Jan to 15 April). Figure 2 shows the peak of total cases may happen in mid of April.

**Figure 2.**
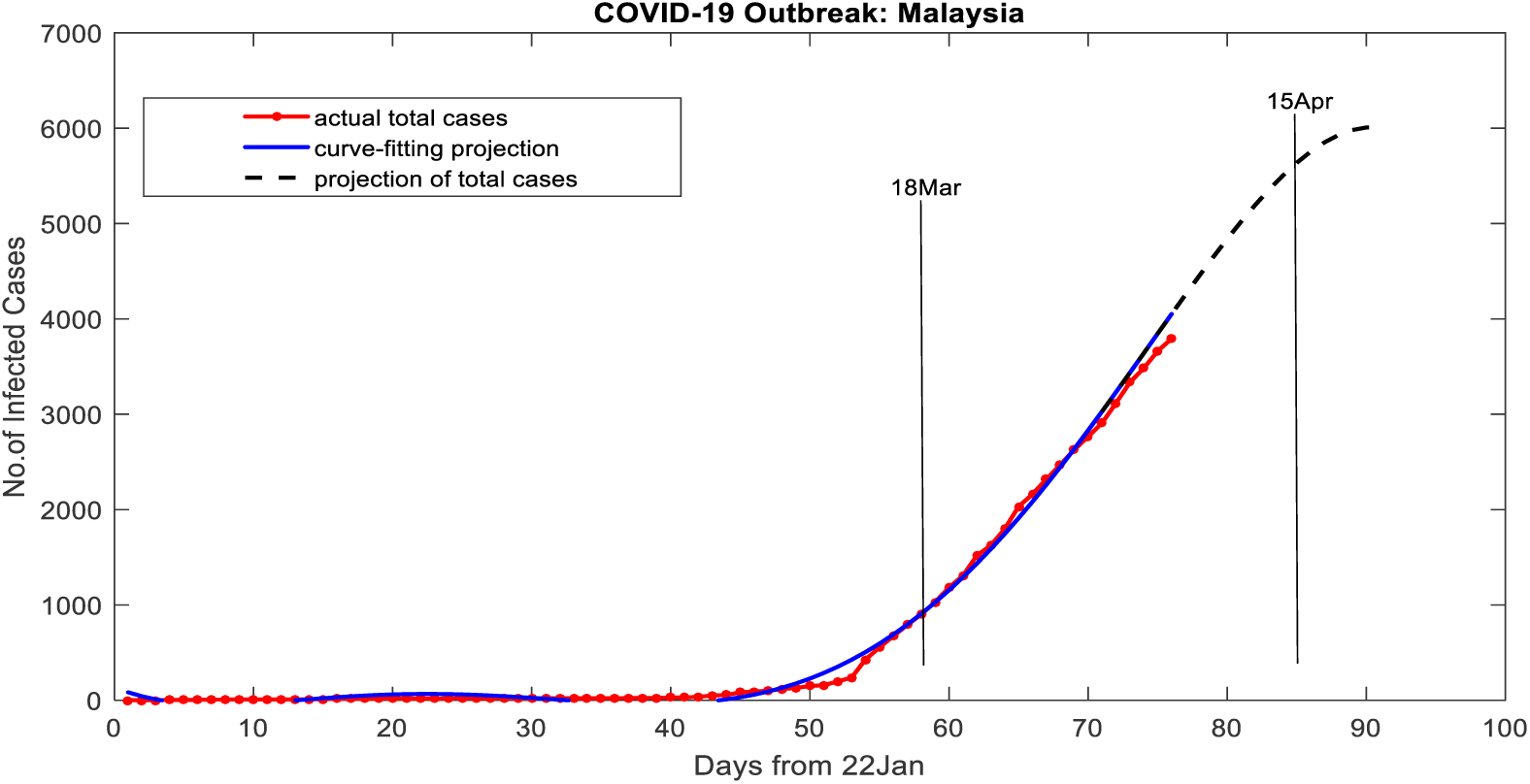
Projection number of Covid-19 infected cases in Malaysia by 15 April 2020

#### 3.1.2. Phase 2: Projection of recovery period to model flattening of the curve

The projection of COVID-19 recovery period can be studied from patterns and trends of various countries. From the observation, it was found that the recovery cases from China and Korea can be used as a basis for extracting valuable information. China took around 25 to 30 days to flatten the curve of total cases while Korea took around 15 to 20 days to flatten the total cases, as shown in Figure 3. Considering a moderate initiative, enforcement, people reaction and behavioral towards Coronavirus pandemic in Malaysia, 28 days of recovery period is quite reasonable to be applied as a conservative approximation for Malaysia. This also supported by 14 days of incubation periods for respiratory recovery for coronavirus patients. The minimum control order (MCO) of 14 days is also being considered to support the assumption.

**Figure 3.**
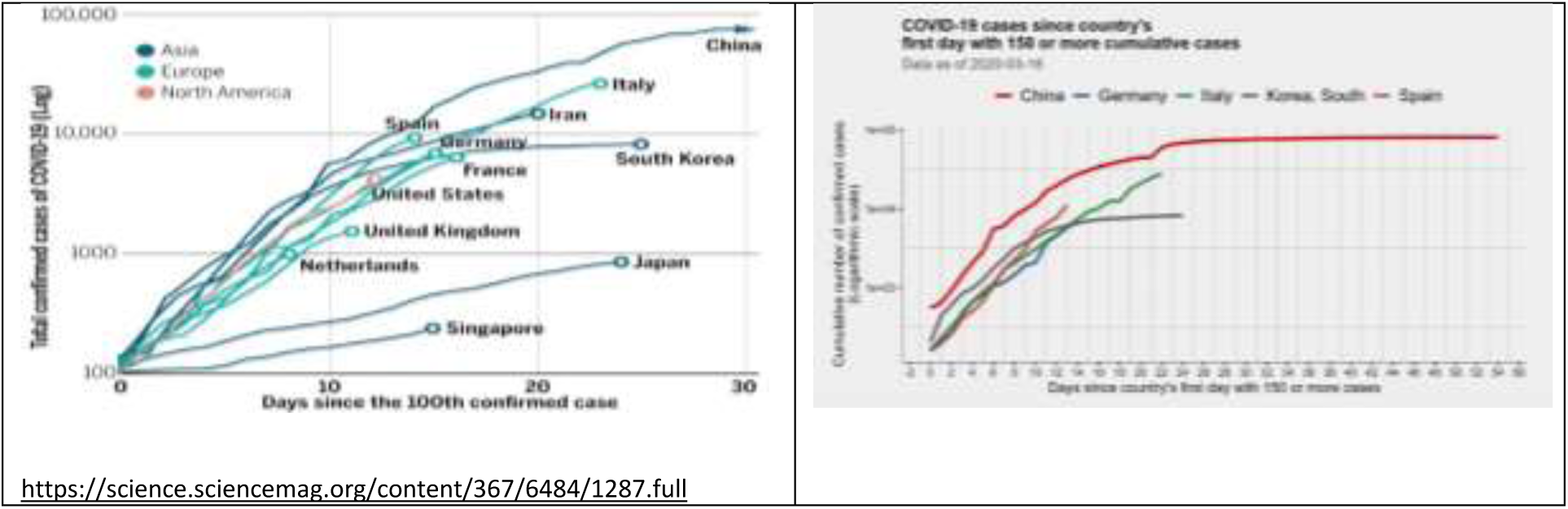
COVID-19 cases from other countries

Statistical method based on probability density function (PDF) is applied to fit and match the increasing rate, peak value, steady-state value, and declining rate of the active cases. For initial projection, consider the trend is similar to a symmetrical bell shape of a normal distribution pattern (also known as Gaussian distribution) to forecast the recovery curve of the active cases. The general equation of Gaussian distribution is given by the following equation.

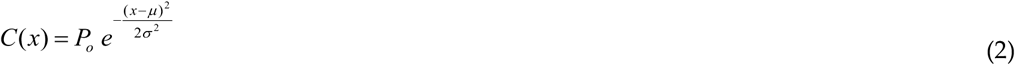

The goodness of fitting to actual data of active cases is performed by simulating various values of mean *μ*, standard deviation *σ* and constant *P*_*o*_ as a function of days *x*. For a recovery projection, active cases (i.e., total cases minus recovered cases) is plotted instead of cumulative of total infected cases. This is to explain why the peak value of recovery plot is lower than peak value of total infected cases.

Figure 4 shows the results of the prediction. It is expected that the active cases reaches its peak in the middle of April. The active cases may end by end of May, on condition of full compliance of the MOC.

**Figure 4.**
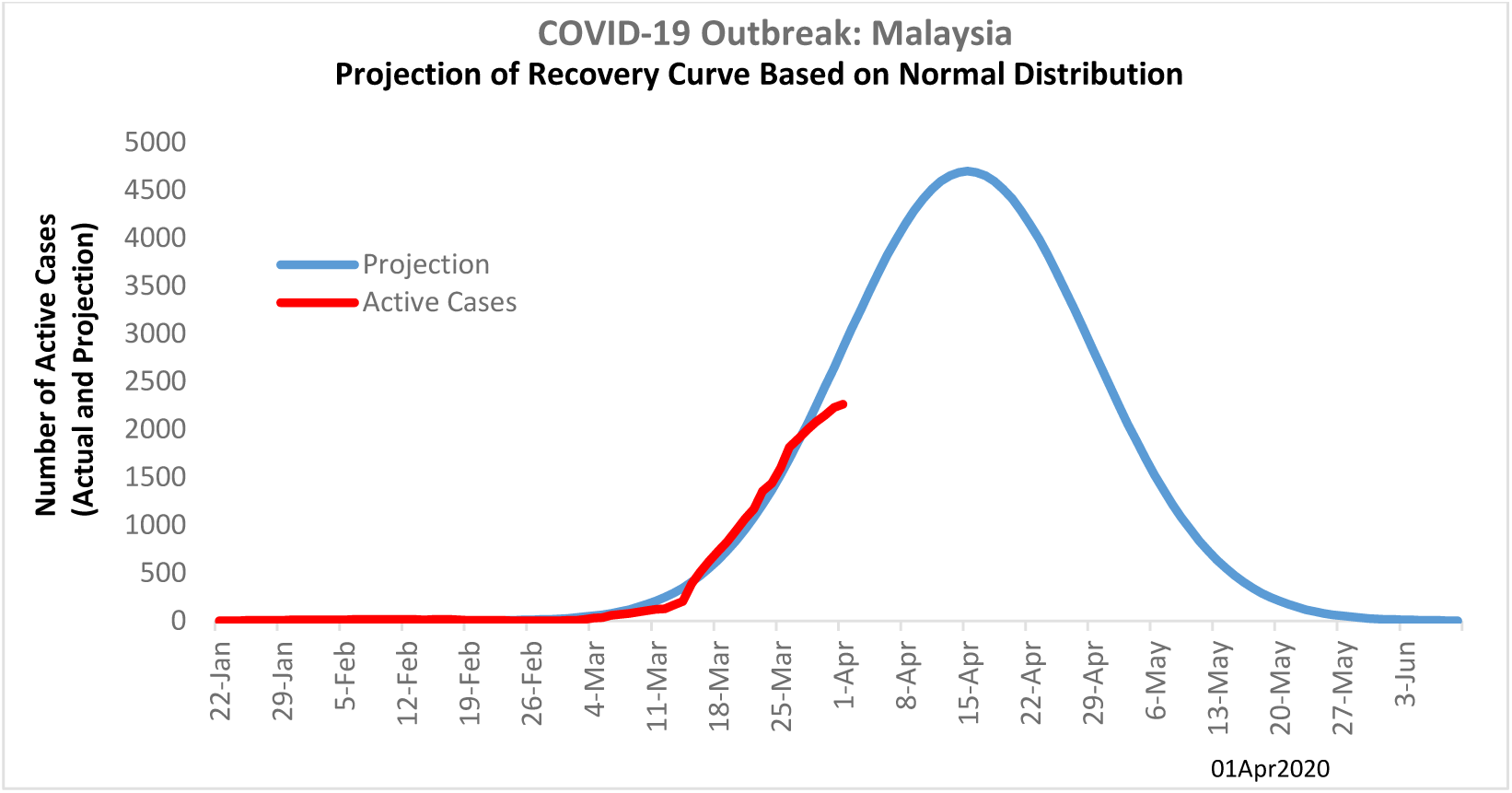
Recovery projection of active cases for Malaysia

It is a trivial solution that the curve of *C*(*x*) is symmetry before and after the peak, however, it may be true all the time especially when dealing with uncertainties. Some uncertainties may happen, such as capacity of healthcare system, social distancing and compliance to MCO in. Figure 5 show the unsymmetrical curves of active cases in China and Korea.

**Figure 5.**
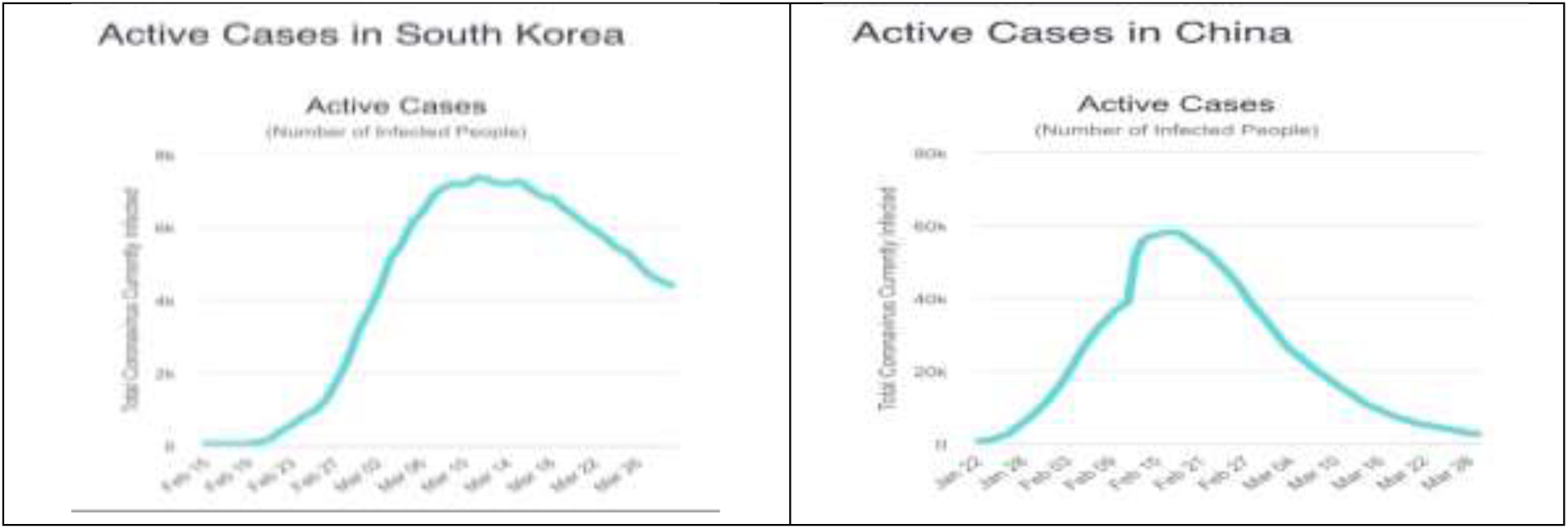
Projection of recovery trends of active cases in Korea and China with skewness effect [23].

The unsymmetrical of the curves can be model using skewness effect to both sides of the distribution curve (i.e., before and after the peak value). In this case the skewness effect is applied on the right side of the curve because it is most likely people may not fully comply with MCO rules and regulations (social distancing, crowd gathering and etc.) after long duration of MCO. The skewness effect is modelled by varying standard deviation on the right side (after the peak value) of the normal distribution.

The degree of compliance can be correlated with standard deviation. For example, the purple line in Figure 6 shows the skewness of 68% with confident level of 3*σ* (three times standard deviation), where in this case it is defined as a “moderate compliance”. It is expected that people will maintain high percentage of compliance (more than 90%) during MCO and pay more attention to it during the declining period of the active cases.

**Figure 6.**
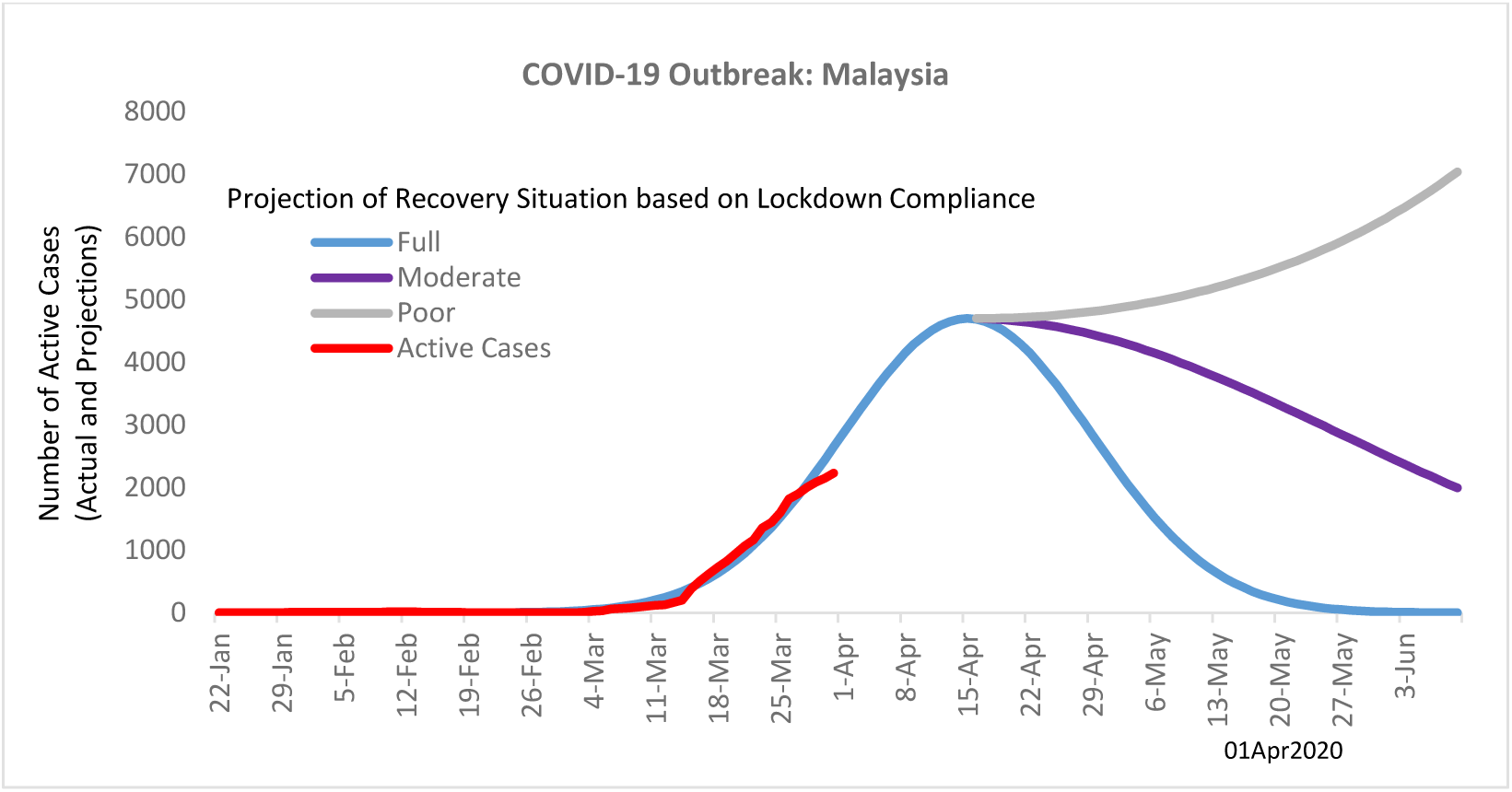
Projection of recovery trend for Covid-19 cases in Malaysia

The high increment of cases (i.e., big jump) from 238 to 428 cases in a day between 14^th^ March 2020 and 15^th^ March 2020 made a huge difference in the next twelve days. The difference is around 1250 cases, as shown in Figure 7. The degree of compliance during lockdown (MCO) period is crucial.

**Figure 7.**
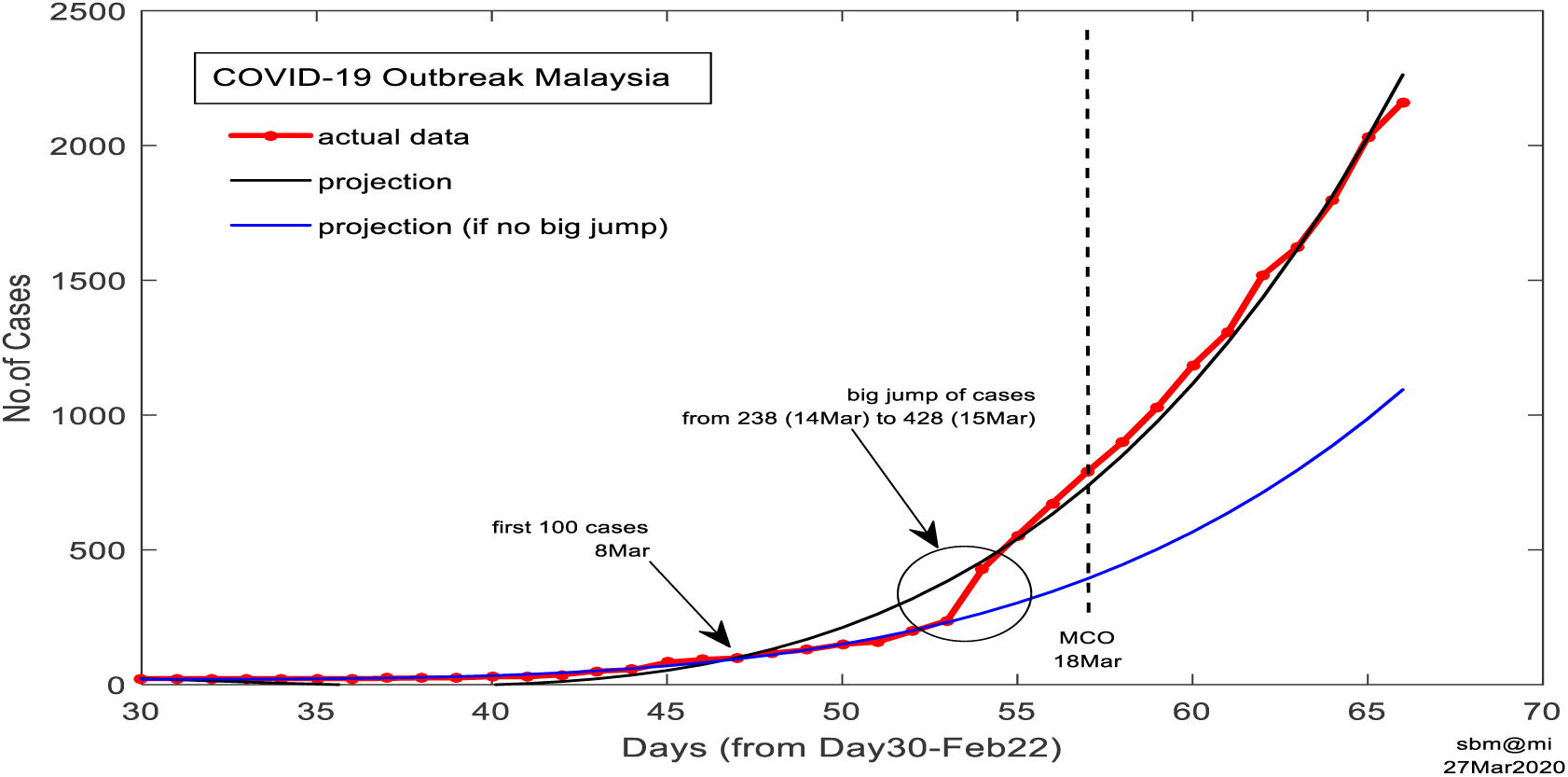
‘Big-jump’ of cases in a single day indicates bad upcoming situation

### 3.2. Method 2: SIR Modeling

The SIR modeling for prediction assumes a fixed reproduction number *R*_*0*_ without considering any interventions. Thus, if the training data to develop SIR model is taken prior to any health intervention such as Movement Control Order (MCO), the prediction of future outbreak would be based on the estimated parameters from the training data. Basically, the basic reproduction number, *R*_*0*_ reflects the ration of the infection from one infected patient to others.

Figure 8 shows the simple illustration of SIR model, where there are two parameters β and γ. In general, the SIR model is used to model how much of the susceptible population get infected through exposure in the rate of parameter β and how the infected population eventually recovered or die in a rate of γ. The equation of the SIR model used in this study is as follows:

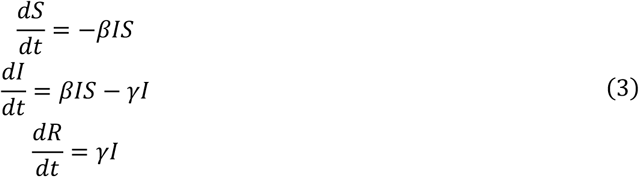

**Figure 8.**
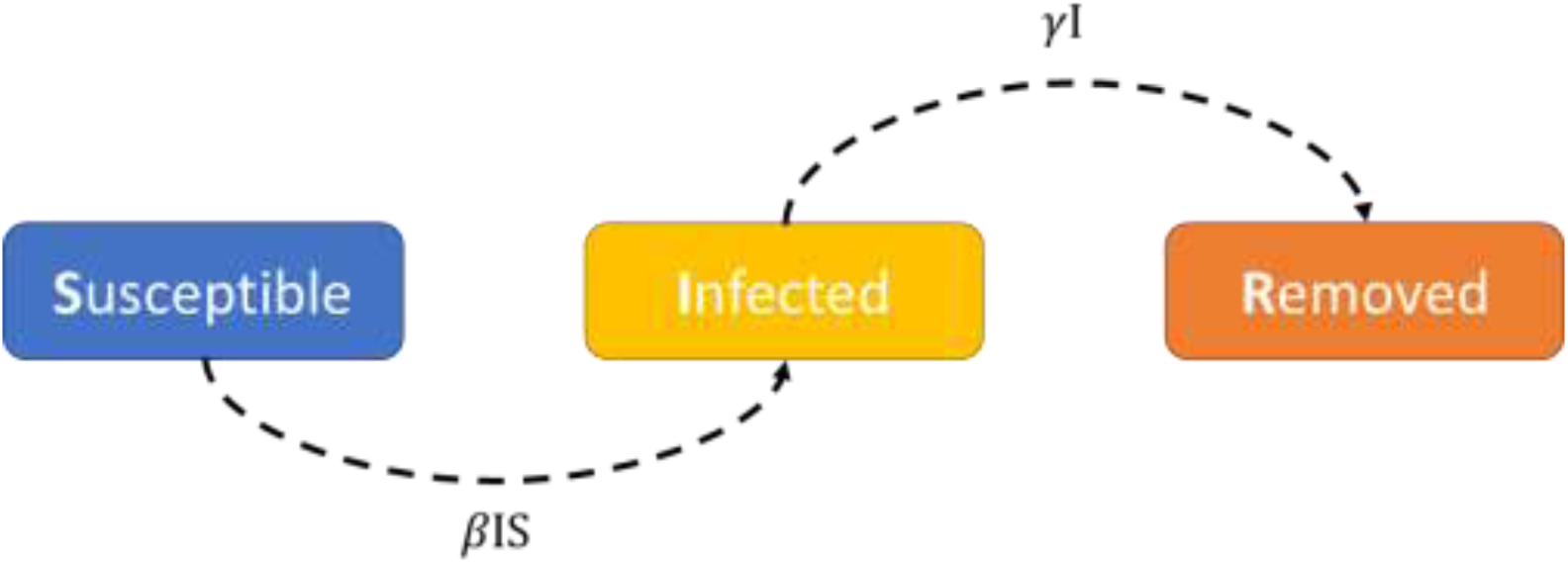
A simple illustration of SIR model

**Figure 9.**
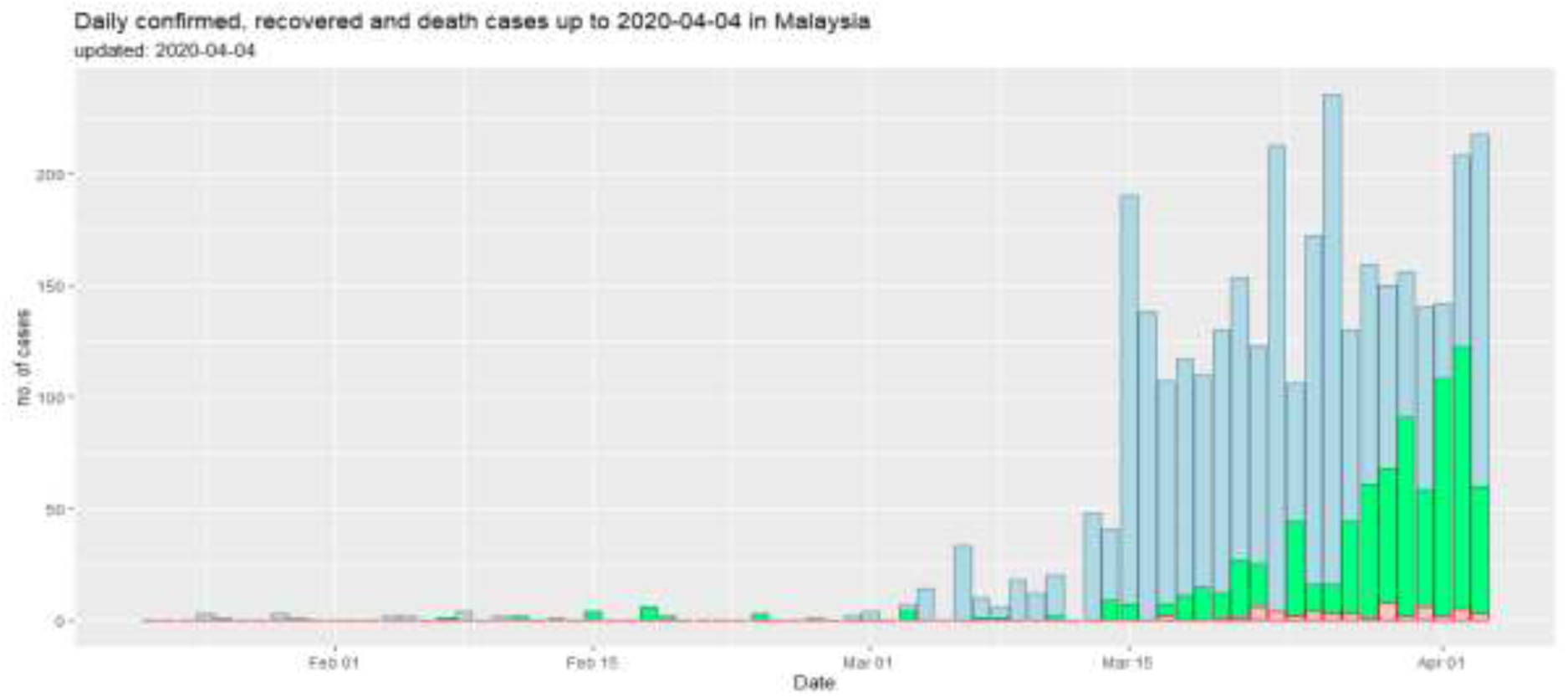
Daily confirmed (blue), recovered (green), and death (red) cases in Malaysia recorded since 22^nd^ January 2020

**Figure 10.**
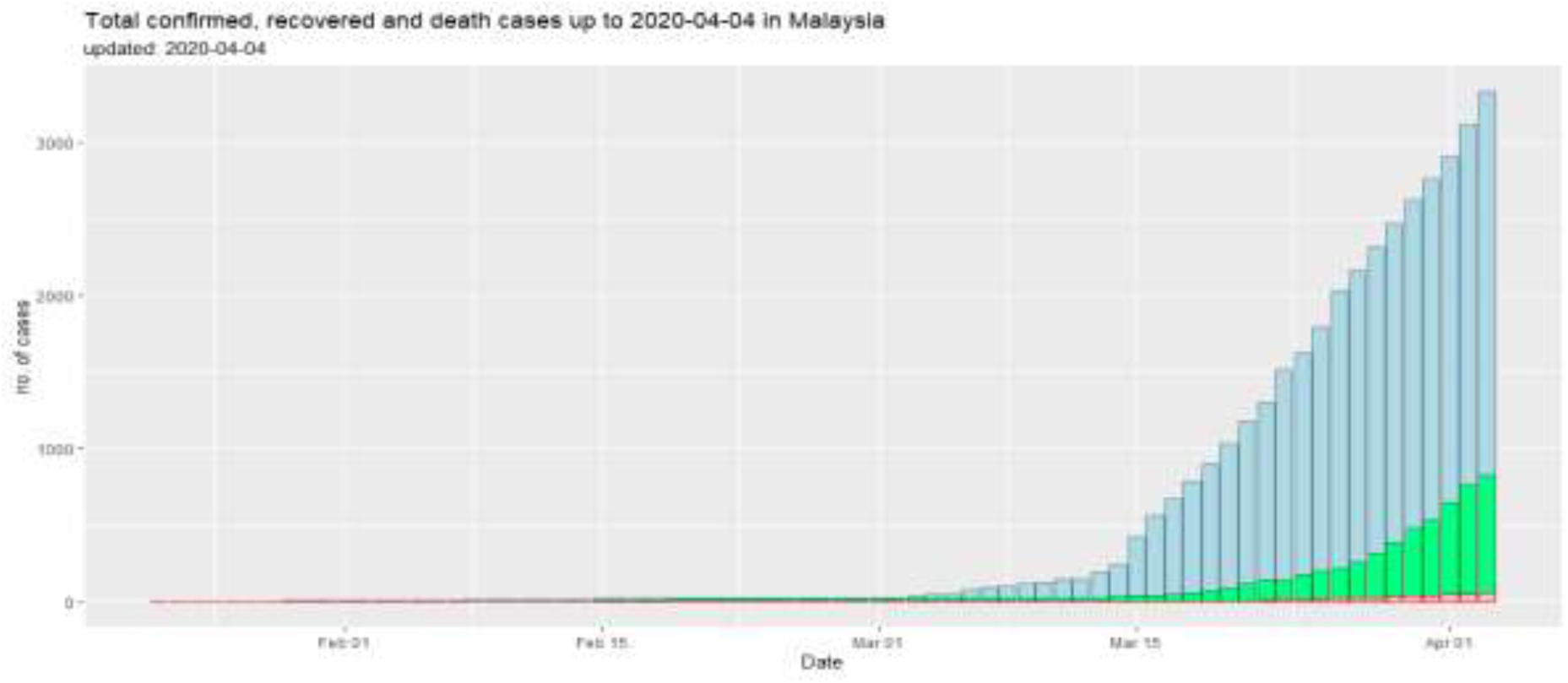
Cumulative confirmed (blue), recovered (green), and death (red) cases in Malaysia recorded since 22^nd^ January 2020

Equation 3 shows the rates for each of the components in the SIR model at a time. The model we implemented does not consider the effect of natural death or birth rate with the assumption of the outstanding period of the disease is shorter than the lifetime of human. The first part of the equation is corresponding to the susceptible (*S*) where 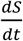 represents the population of *S* at time *t*. Assuming at a time *t*, the probability of the susceptible to get infected is the fraction of susceptible over the total population considered in the modeling, 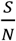 and the probability of the infected individual to infect the susceptible is the fraction of the infected (*I*) at time *t*, over the total population in the model, 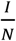. Assume that eventually the portion of the susceptible will be reduced whenever an individual gets infected. Thus, a negative of rate of transmission, β, is included in the formula. On the other hand, the second part of the equation represent the changes in the infected (*I*) over time. Similar concept with the first part of the equation, assuming the infected individuals are increasing with the transmission rate of β and at the same time, there will be a portion of infected individuals who healed from the disease or died of it with a rate of γ. The last part of the SIR model represents the removed population take account the individuals who recovered from the disease or those who died. This number of this R population will eventually be increased with the rate of γ.

#### 3.2.1 Current Malaysia COVID-19 statistics

Malaysia currently (as of 3^rd^ April 2020) has recorded a total incident cases of 3333 with 827 have recovered and 53 death cases. Figure 2. shows the overview of the reported confirmed cases, recovered cases, and death cases of Malaysia from 22^nd^ January 2020 while Figure 3 shows the cumulative cases within the same period.

Based on the data, the number of confirmed cases in Malaysia still grows exponentially. In terms of the growth rate of the confirmed cases, we analyze the data from 1^st^ March 2020 until 3^rd^ April 2020, and the average of the growth rate is 1.16. The data is shown in Figure 11.

**Figure 11.**
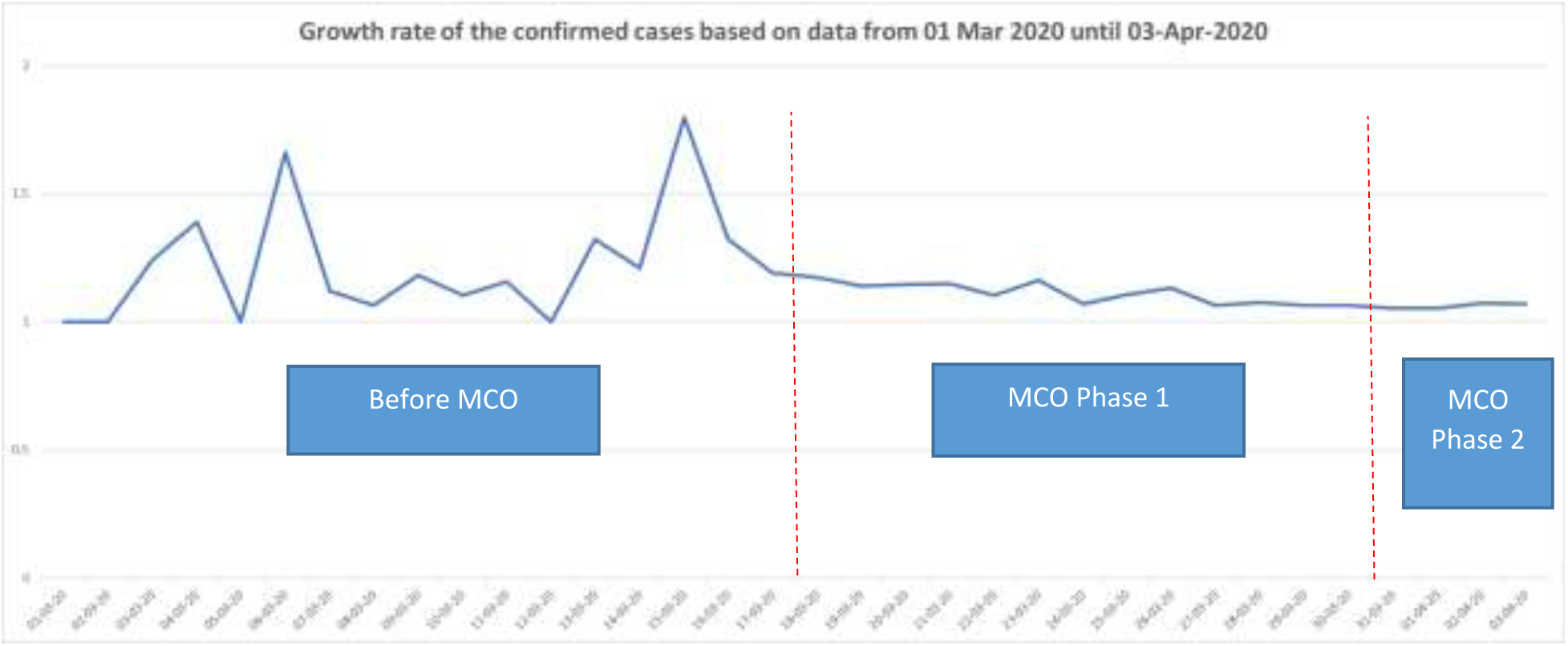
Growth rate of the confirmed cases in Malaysia.

Based on Figure 11, it clearly shows that few days before the implementation of the Movement Control Order (MCO) has cause drastic increase of the confirmed cases (up to 80%), and the growth rate has dropped but remaining considerably high or exponential for the rest of the days within the MCO period. This also indicate that the implementation of the MCO is effective to reduce sudden spike of the growth of the confirmed cases of COVID19.

In terms of the case fatality rate, based on the data up to 03 April 2020, the cumulative fatality rate of the COVID19 in Malaysia is 1.67% with the highest daily case fatality rate of 5.33% (see Figure 12).

**Figure 12.**
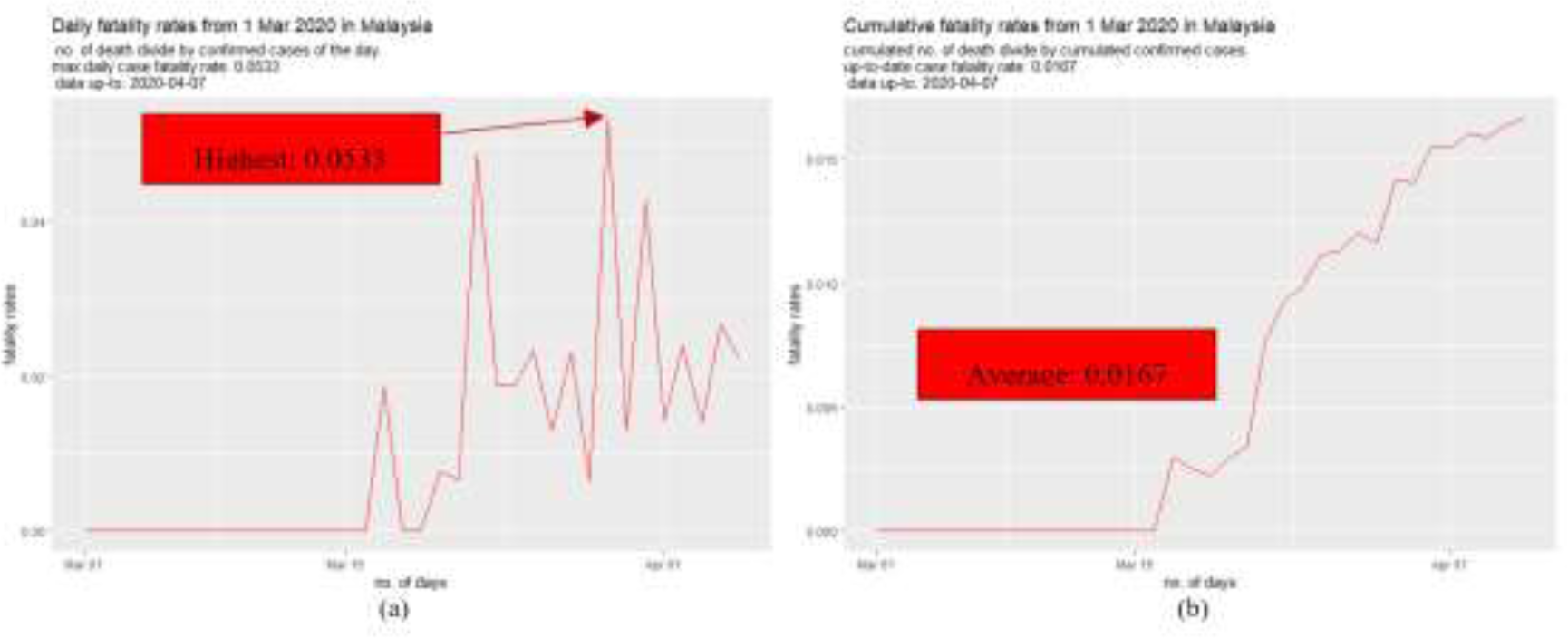
(a) Daily case fatality rate and (b) cumulative fatality rate of COVID19 in Malaysia.

Meanwhile the recovery rate of COVID19 in Malaysia has shown the sign of increasing trend for the past week, with the max recovery rate recorded of 76.06% (122 cases) in one day. The cumulated recovery rate of COVID19 in Malaysia currently is 27.44% (refer to Figure 13).

**Figure 13.**
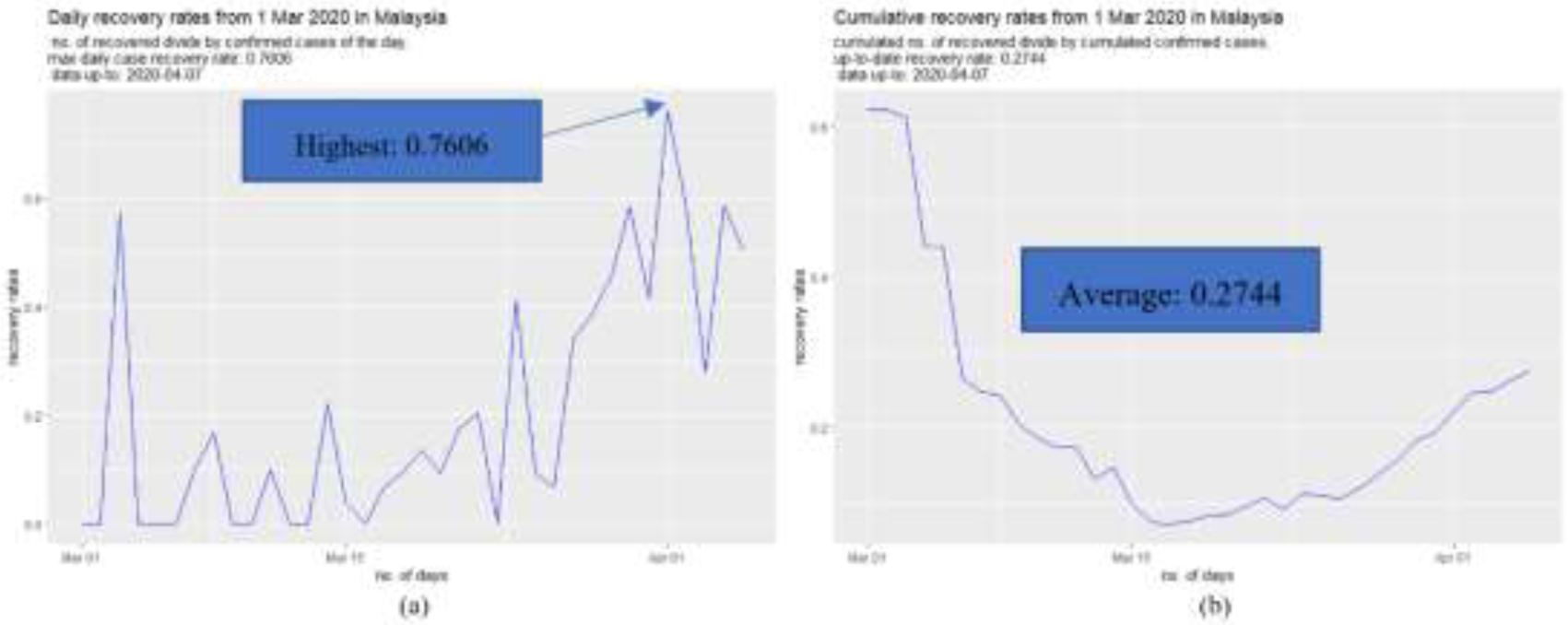
(a) Daily recovery rate and (b) cumulative recovery rate of COVID19 in Malaysia.

#### 3.2.3. Results

In our study, SIR models were generated with two different sets of data. First SIR model was generated using the data obtained before the MCO period (we called it non-MCO) which is 1st March to 17th March 2020. Meanwhile, second SIR model was generated using the data obtained during MCO phase 1 period (we call it MCO) which is 18th to 31st March 2020. During the first phase of the experiment, these two SIR models (non-MCO) and (MCO) assume that the total Malaysian population of 32.6 million (which is 100%) is susceptible to be infected. Meanwhile the second phase of the experiments, we assume only 30% of total Malaysian population is susceptible to be infected.

Figure 14 shows the predicted cases using the SIR model (non-MCO with 100% population). The model peaks at 2.6 million cases, which is equivalent to 8% of the total Malaysian population on 20th April 2020. However, second SIR model (MCO with 100% population) shown in red, shows a lower peak (at 630K predicted cases) on 31st May 2020.

**Figure 14.**
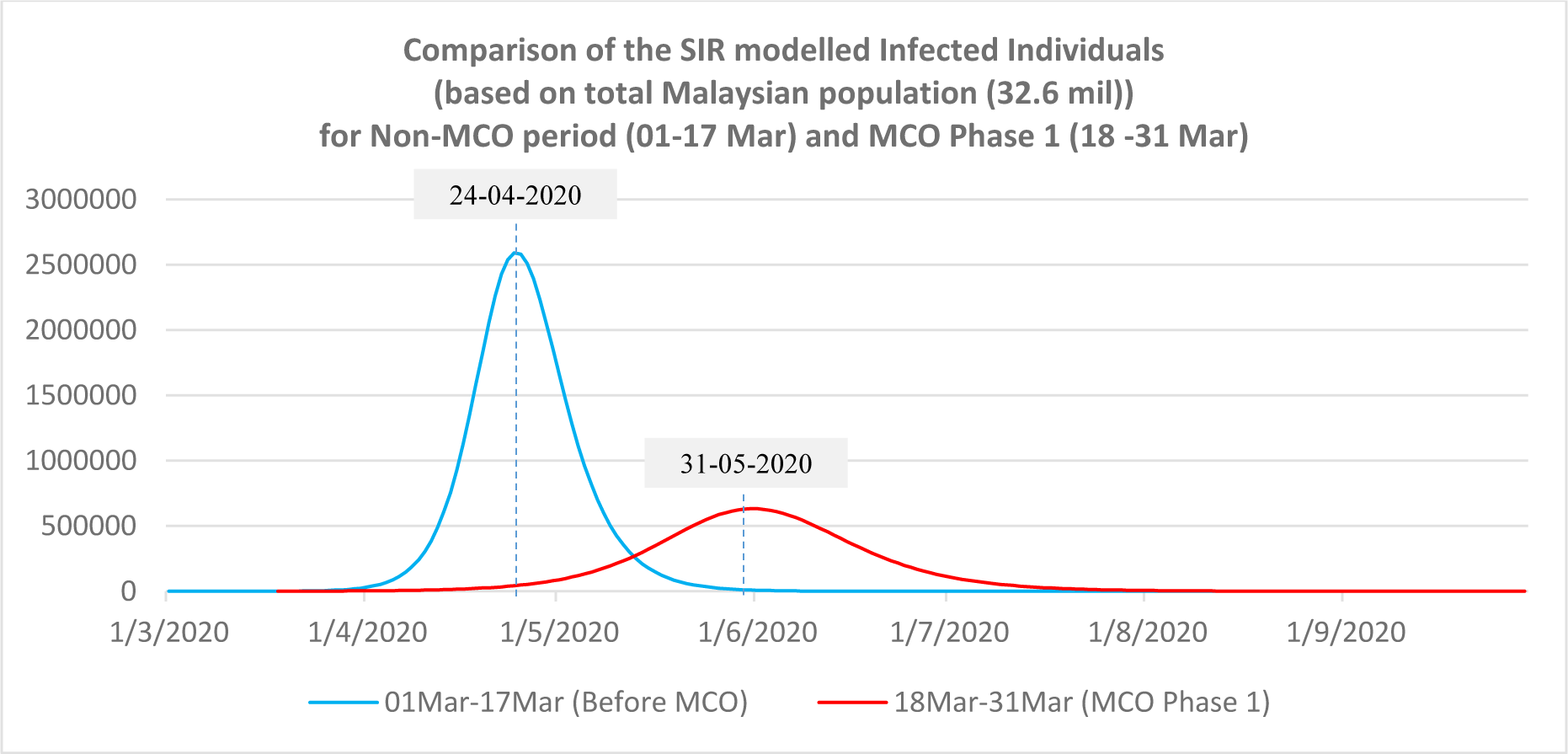
Comparison of the SIR modelled infected individuals (based on total Malaysian population).

In general, this graph illustrates the impact of MCO implementation which has started on18th March 2020. Based on the comparison with the actual reported cases up to 3rd April 2020, there is a total difference of 50k cases between actual and the projected number of cases by the first SIR model (non-MCO with 100% population - see Figure 15). On the other hand, the second SIR model (MCO with 100% population) shows a better prediction. A difference of 951 is observed on 3rd of April between predicted and actual cases.

**Figure 15.**
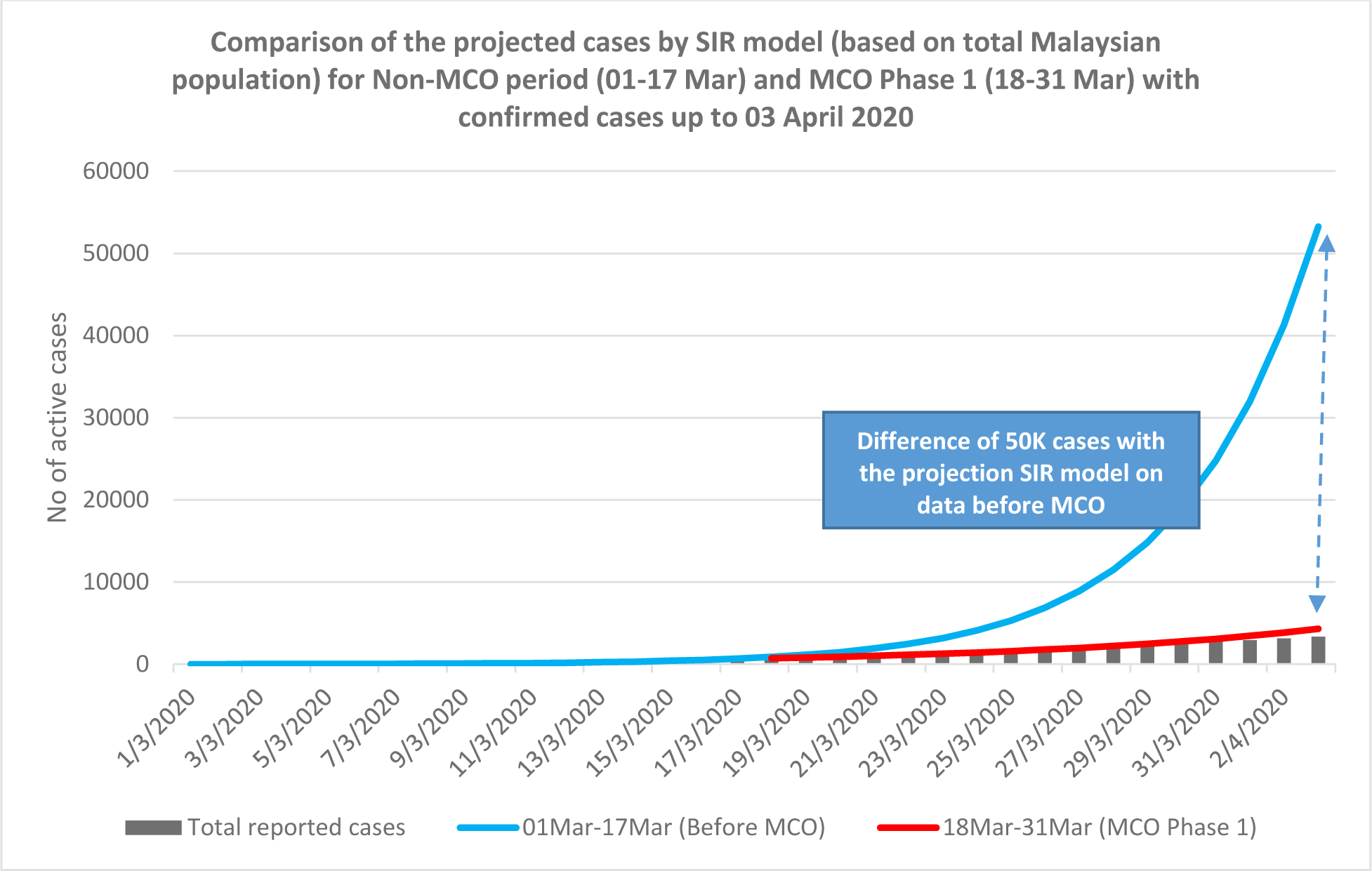
Comparison of the projected cases by the SIR models with reported confirmed case.

In both SIR models (non-MCO and MCO with 100% population), the parameters β and γ are generated through constraint optimization. The estimated parameters (β and γ) for both SIR models are used to calculate the Basic Reproduction Number (R0) by dividing the β with γ. The parameters generated by both SIR models are shown in Table 1.

**Table 1.**
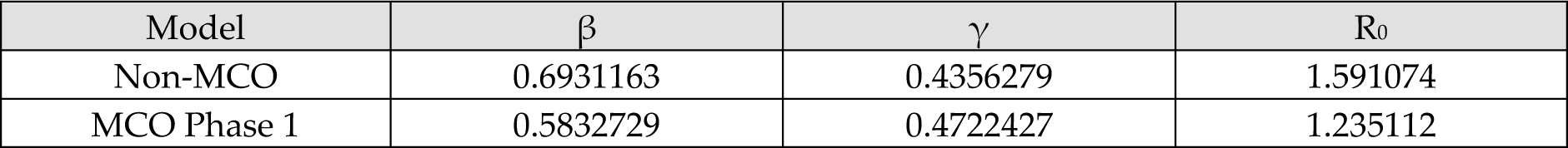
Parameters generated by both SIR models using 100% of Malaysian population

During the second phase of our experiment, we assume that social distancing is practised and 70% of the population adheres to the government order to avoid or minimize contact with the infected patients. Therefore, 30% of the total Malaysian population are assumed to be susceptible to be infected. Using 30% population as susceptible, we produce the following model shown in Figure 16

**Figure 16.**
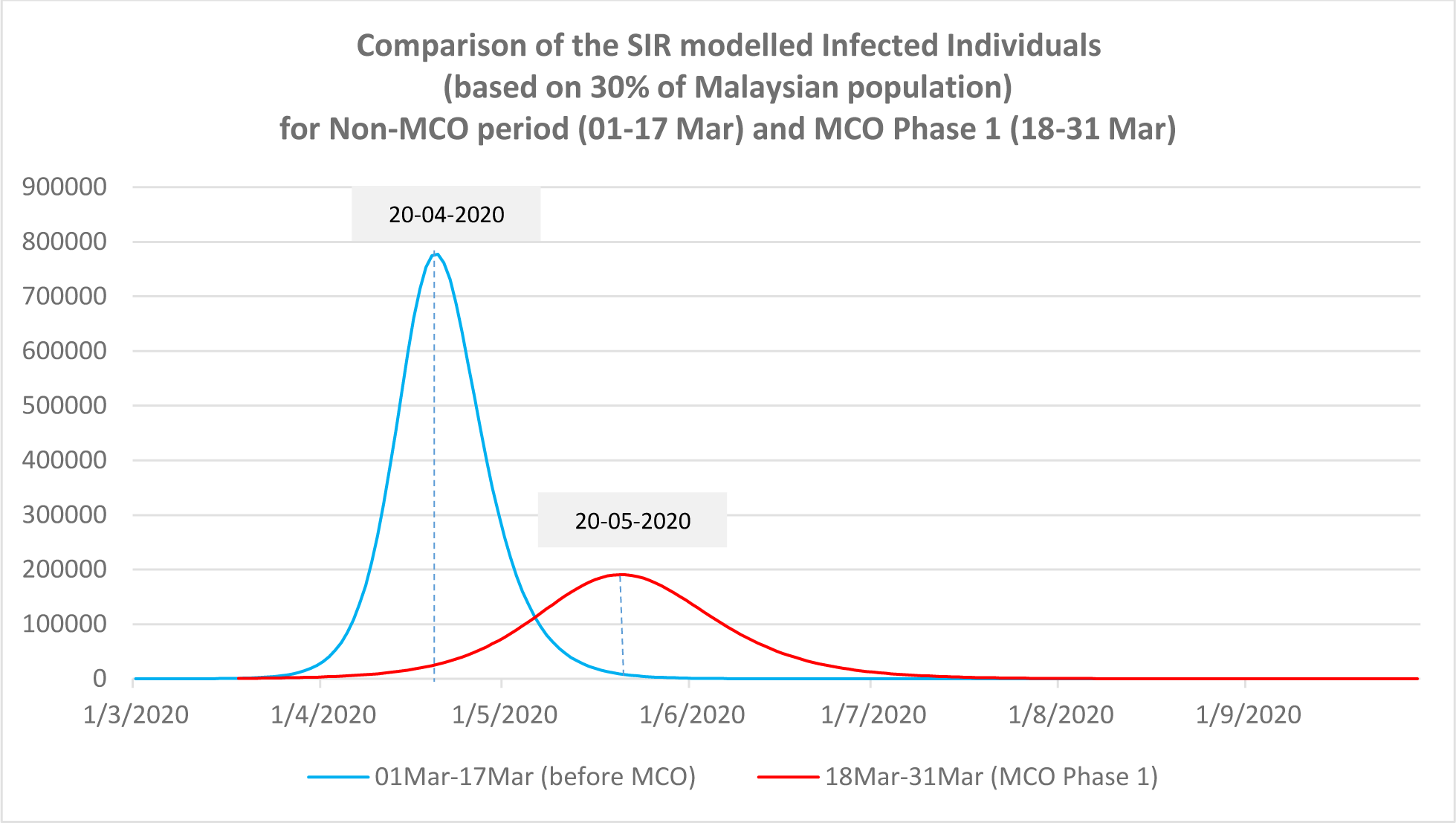
Comparison of the SIR modelled Infected individuals based on 30% of Malaysian population.

Compared to Figure 14, the estimated peak based on SIR model (non-MCO with 30% population) occurs earlier which is around mid of April 2020 (20th April 2020) with predicted max cases near to 800k. While for the SIR model (MCO with 30% population), the estimated peak happens around mid-May (20th May 2020) with about 200k cases. This implies a 75% reduction of cases. Our finding, if the susceptible population is reduced due to social distancing and MCO, the expected peak of cases could be flattened and it will peak at earlier date (around 10 days earlier than the peak of SIR model (MCO with 100% population) as shown in Figure 14). The values of estimated parameters for these models are as shown in Table 2.

**Table 2.** Parameters generated by both SIR models based on 30% population of Malaysia

| Model | $\beta$ | $\gamma$ | $R_0$ |
| --- | --- | --- | --- |
| Non-MCO (30%) | 0.6931304 | 0.4356232 | 1.591124 |
| MCO Phase 1 (30%) | 0.5834263 | 0.4721928 | 1.235568 |

It is important to note that the parameters are very similar to the models generated based on 100% total Malaysian population. The only difference is the time when it peaks and the the maximum number of predicted cases. The projected values of S, I and R are also shown in Figure 17. Figure 18 and 19 show the projected number of Susceptible (S), Infected (I) and Recovered (R) persons based on the SIR models (non-MCO and MCO) using 30% of Malaysian population and 100% Malaysia population, respectively.

**Figure 17.**
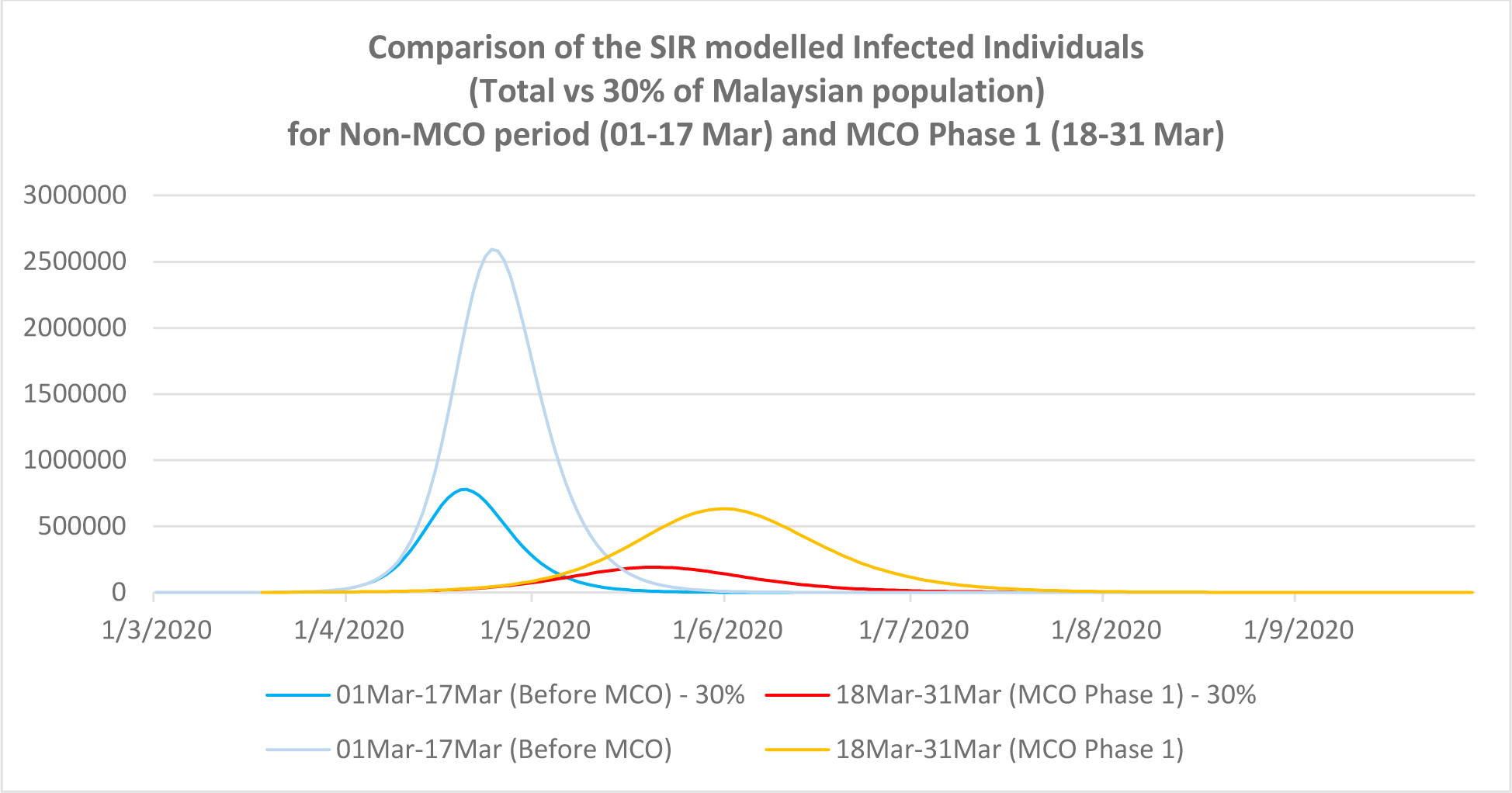
Comparison of SIR model infected individuals based on total and 30% of Malaysian population.

**Figure 18.**
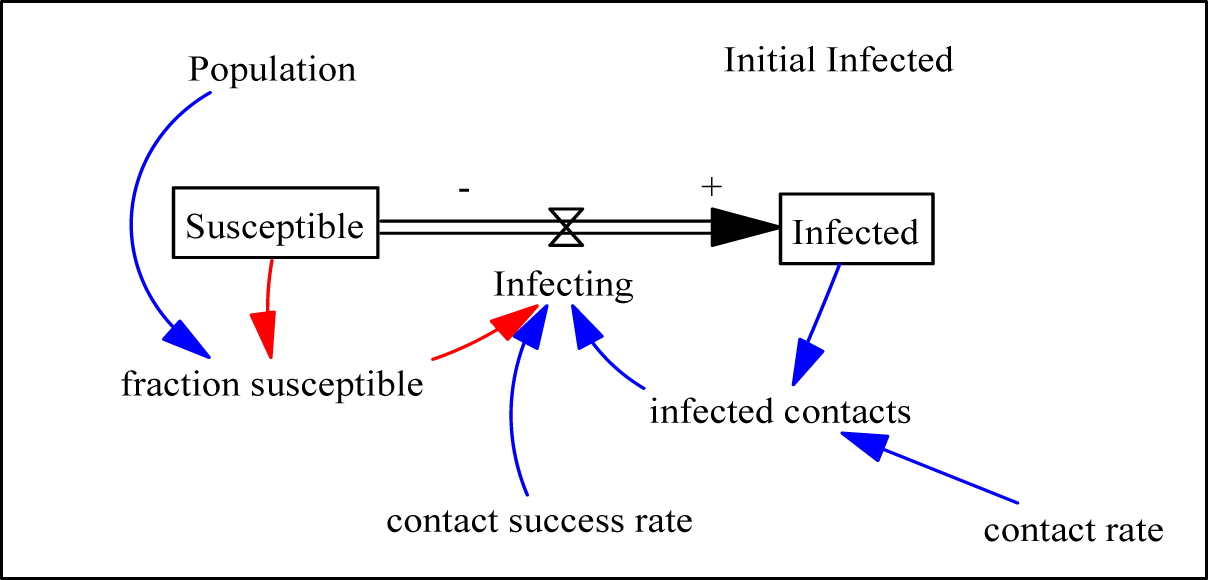
A System Dynamics model for infected cases

**Figure 19.**
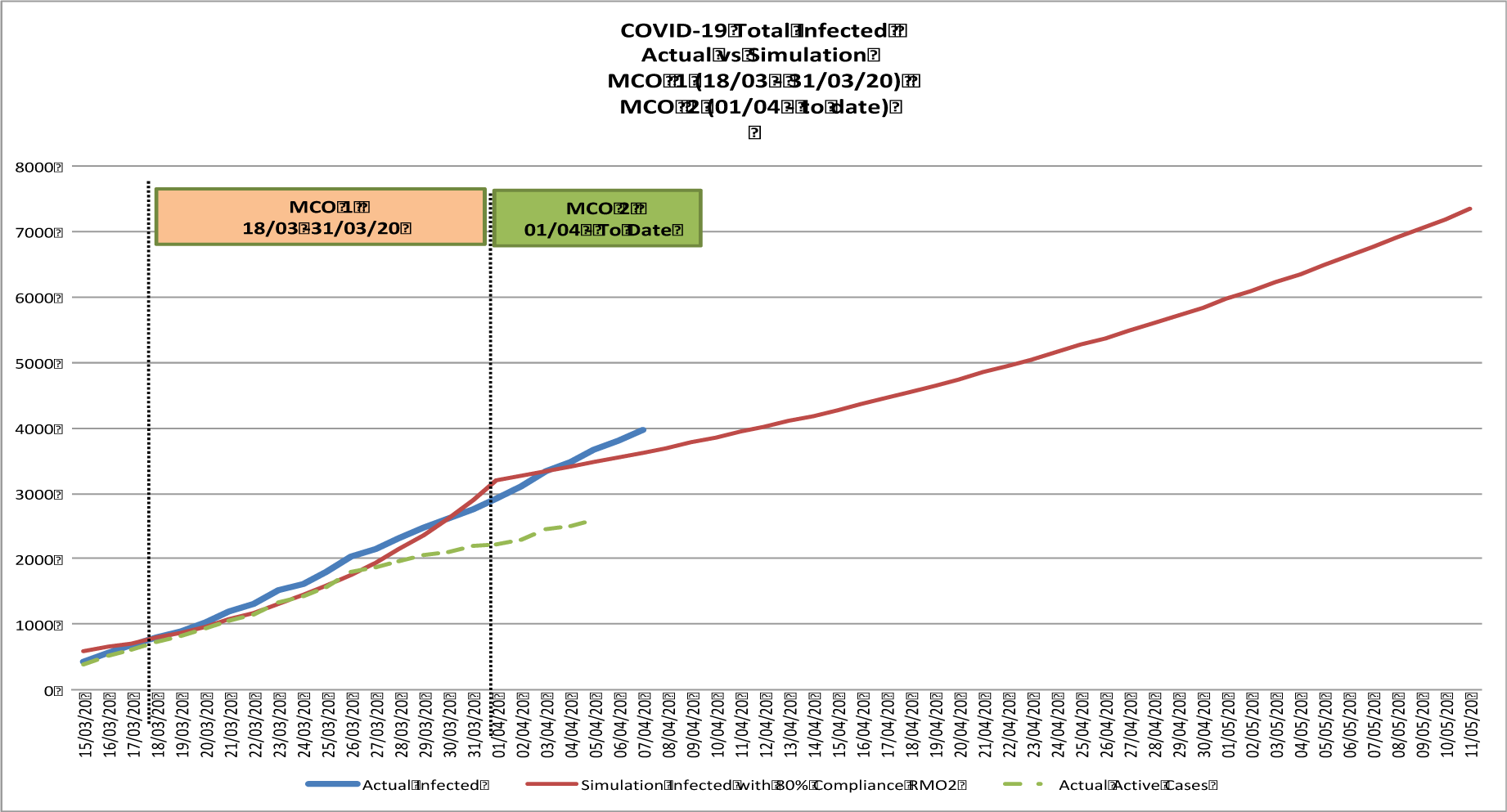
Total infected persons actual vs. simulated output. Active cases are those still infected (actual infected – recovered-death)

### 3.4. Method 3: Systems Dynamics Modeling

Here, we designed a simple system dynamic model to represent the susceptible and infected population by using contact rate, and contact success rate as parameters to understand the effect of Movement Control Order (MCO) Phase 1 and Phase 2 on the number of total infection of the COVID- 19 cases in Malaysia.

#### 3.4.1 Phase 1: Identification of contact rate and contact success rate

A system dynamics model by system dynamics society [28] is utilized by using Malaysian data to understand the current trend of COVID-19 infection under MCO Phase1 and Phase 2. In this model, as shown in Figure 18, the susceptible and the infected population during this COVID-19 outbreak are used to understand the trend. Susceptible population will become infected when in contact with infected person. Daily total infected data are utilized starting from the 1st February 2020 with initial total infected of 8 persons as the value in our simulation. For the purpose of the simulation, value for the contact rate and contact success rate are required to replicate the total infected trend in Malaysia.

Contact rate represents the number of people an infected person has contact with per day while contact success rate is success rate an infected person infect other people. Since COVID-19 can be transferred through droplets of the infected person when coughing or sneezing which can go as far as 1 meter, 5 people is used as the contact rate. This estimate is to represent that within 1 meter radius, there can be an average of 6.5 persons in contact with the infected person in that very close proximity. Thus, 5 people is estimated to be a reasonable value to start with. However, those who become infected by touching surfaces contaminated with the droplets of infected person is difficult to model, hence is not included in our simulation.

WHO has estimated the average number of people successfully infected by one infected person parameter (R_0_,) of 1.4 – 2.5 for COVID-19, which means that an infected person can successfully infect 1.4 – 2.5 other people in average. In our model, R_0_ is translated into contact success rate of 0.021 or 1 infected person can successfully infect other 2.1 people, which is still within the estimate by WHO.

Important parameters used in the model are shown in Table 3.

**Table 3.** Parameters of the SD model

| Parameters | Description | Value |
| --- | --- | --- |
| Population | Malaysia Population as per 2019 | 32,680,000<br>( https://www.dosm.gov.my/) |
| Initial Infected | Total number infected as per 01/02/20 | 8<br>(https://www.worldometers.info/coronavirus/country/malaysia/) |
| Contact Rate | Number of people an infected person has contact with per day | 5<br>(this value projects the simulation to match actual data) |
| Contact Success Rate | A fraction to show infection rate reflecting $R_0$, the average number of people successfully infected by one infected person | 0.021<br>(This equals to $R_0 = 2.1$. WHO has estimated in January 2020 that $R_0$ for COVID-19 is 1.4 and 2.5 [29]. However, preliminary study by [30-31]; and Imperial College London estimated $R_0$ to be between 1.5 and 3.5 [32]. This means 1 single infected person in Malaysia can possibly successfully infected another 2.1 person in average. Just to note, $R_0$ for common flu= 1.3, $R_0$ for SARS is 2.0 and outbreak with $R_0$ below 1 will gradually disappear. |
| Susceptible | People who can catch the infection considering Malaysia Population deducted with Initial Infected | Susceptible initial value = population-initial infected (see row below) |
| Infected | Total Infected considering susceptible and recovered | Initial value = Initial infected (as on 1<sup>st</sup> February 2020) |

The first simulation was conducted to find the value of contact success rate and contact rate of the infected person so to match the output from the simulation to actual data during the MCO Phase 1 and Phase 2. Actual data from the start of MCO Phase 1 and MCO Phase 2 was compared with data from the simulation during the same period as shown in Figure 19. Mean Absolute Percentage Error (MAPE) was calculated to estimate the percentage of error in the projection of the simulation output. Using the following formula:

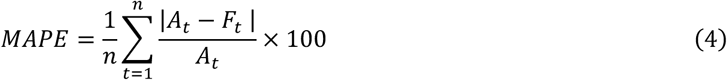

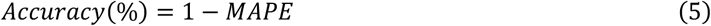

Data for actual total infected from 18^th^ March 2020 to 6^th^ April 2020 was used as benchmark for the projection as this is the duration with best fit of actual data and simulation output. The result of the MAPE calculation is as following table:

Results from the simulation in Figure 19 is obtained by modeling the reduction of contact rate of the infected person from 1:5 to 1:1 during MCO2 (refer to Figure 20 where there are more enforcement in comparison to MCO1). Contact rate of infected person during MCO 1 remains at 1:5, which means in MCO1, an infected person still has large number of contacts.

**Figure 20.**
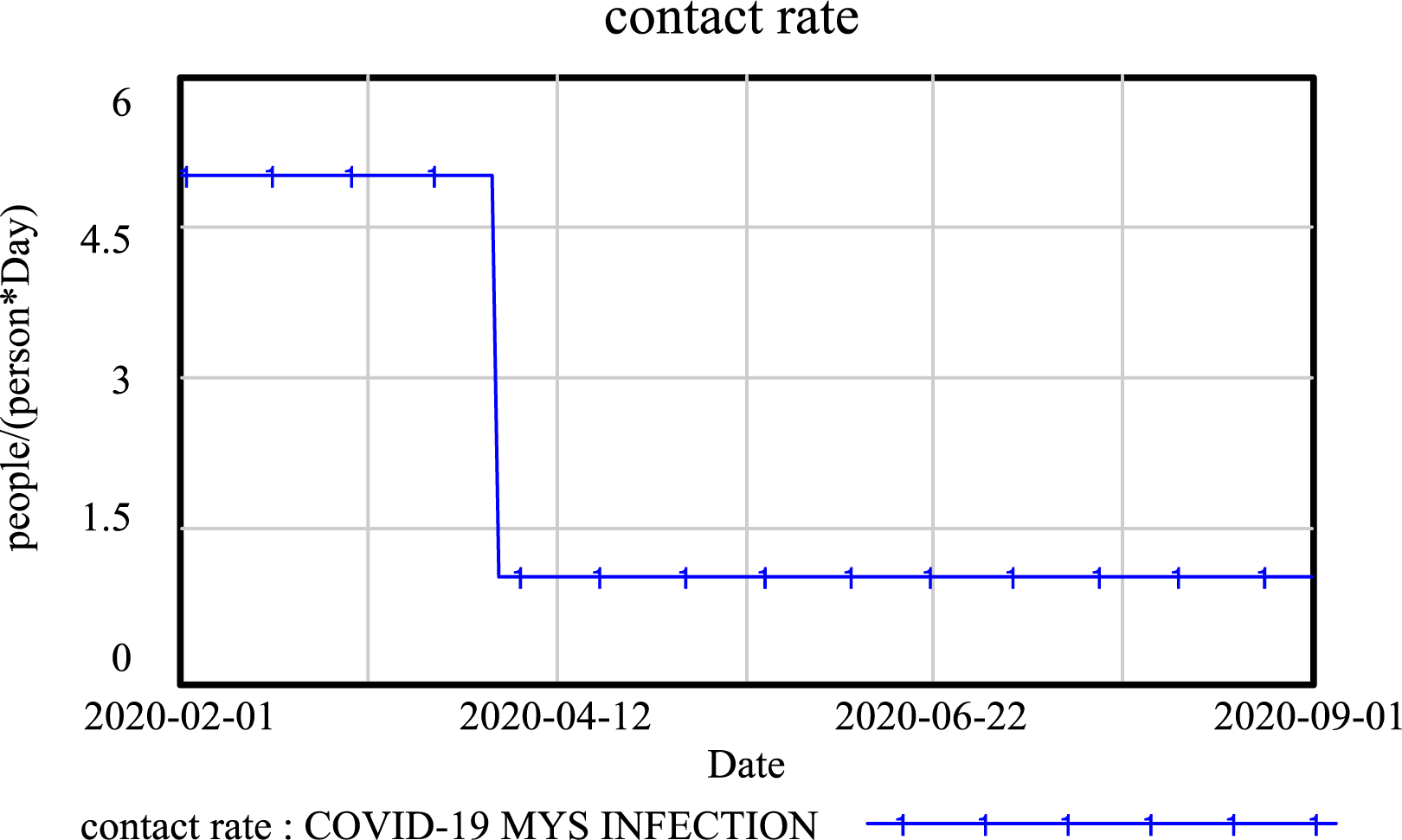
Reduced contact rate from 1:5 to 1:1 per infected person during MCO2

**Figure 21.**
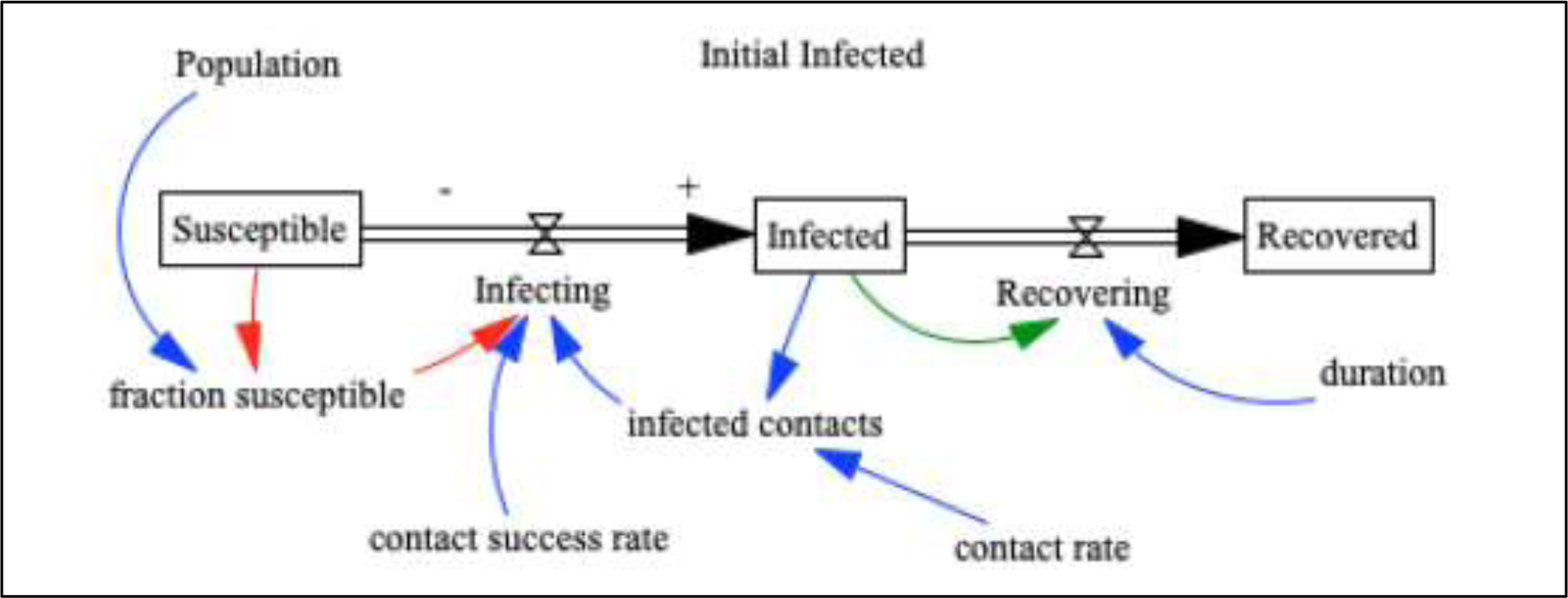
Expanded SIR System Dynamics Model

The model is then expanded to reflect SIR model by including the number of recovered person in order to understand the trend of the active infected cases. The simulation is run from the start of MCO1 (18^th^ March 2020) and the output is compared to the actual data.

In the first wave of the first 22 cases, the average hospitalization period is assumed to be 14 days. However, referring to WHO, mild cases can take up to 2 weeks to recover while more serious cases can take up to 3-6 weeks to recover. Parameters of this expanded model are shown in Table 5.

**Table 4.** Result of MAPE calculation

| Data duration | Parameter | MAPE (%) | Accuracy (%) |
| --- | --- | --- | --- |
| 18/03/20 -06/04/20 | Contact Rate = 5<br>Contact Success Rate:<br>0.021 (R<sub>0</sub>=0.21) | 6.76 | 93.24 |

**Table 5:** Parameter values for expanded SD model

| Parameters | Description | Value |
| --- | --- | --- |
| Initial Infected | Total number infected as per 18/03/20 on the start of the MCO1 period | 790<br>(source: https://www.worldometers.info/coronavirus/country/malaysia/) |
| Contact Rate | Number of people an infected person has contact with per day | 5 reduced to 1 as in Figure 20 (this value projects the simulation to match actual data) |
| Infected | Total Infected considering susceptible and recovered | Initial value=Initial infected |
| Recovered | Total recovered as per 18/03/20 on the start of the MCO1 period | Initial value=60 |
| Duration | Number of days an infected person take to recover | 21 days (this value refers to WHO data, which best match actual trend in Malaysia) |

This expanded model is simulated under 2 scenarios. The first scenario is contact rate of infected person=5 under MCO1 and contact rate=1 under MCO2 to reflect 80% of MCO2 compliance. The second scenario is to reflect that MCO2 has successfully reduced the contact of all infected person to zero by day 10 (10^th^ April 2020) of MCO2. The contact rate=1 is used for the first 10 days and reduced to zero for the last 4 days of the MCO2. This reduced contact rate is shown in Figure 22.

**Figure 22:**
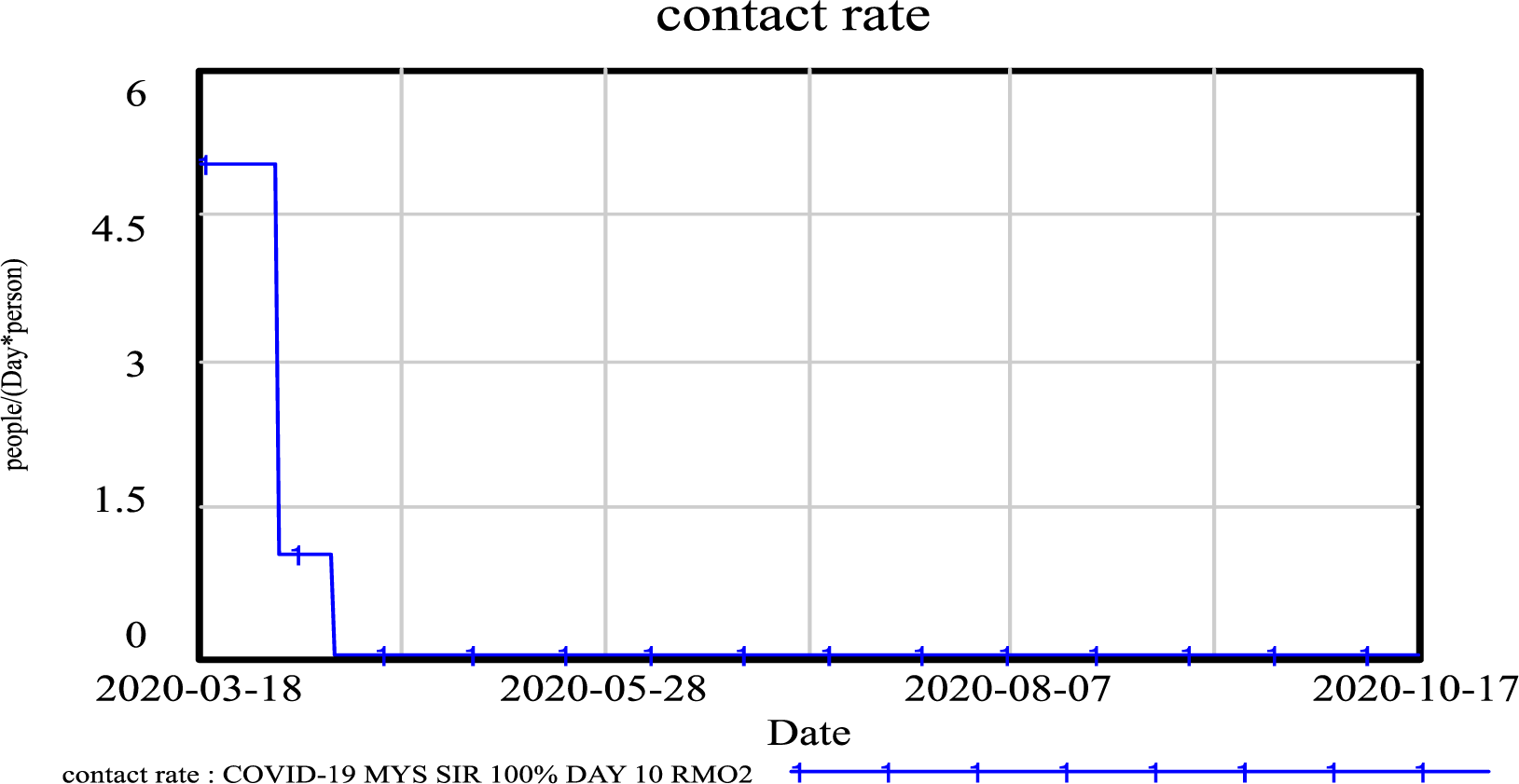
Reduced contact rate of infected person from 1:1 to 1:0 at day 10 MCO2

Output from simulation is then compared to the actual as shown in Figure 23. Based on comparison in Figure 23, actual active infected cases are far higher than the projected active case with MCO2 80% compliance and MCO2 100% compliance from day 10, which means that the MCO is still not complied.

**Figure 23.**
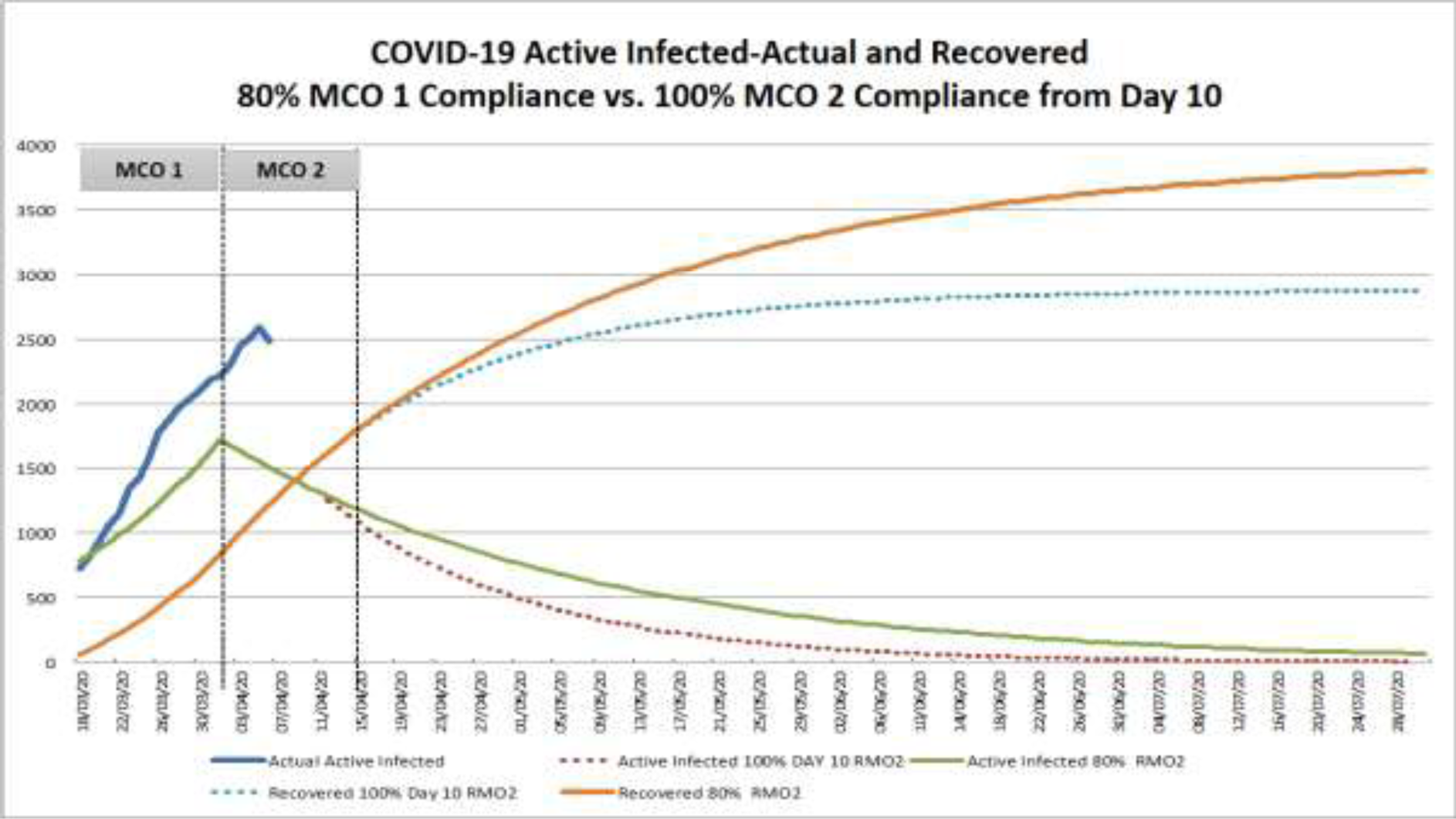
Actual vs. simulated output. Active cases are those still infected (actual infected – recovered- death) under (1) MCO2 80% compliance and (2) MCO2 100% compliance from day 10

A further simulation run is conducted to gain more insights on the trend of the actual cases. The parameter used is *contact rate=5*, which means that an infected person is still estimated having contact with five people. The results from the simulation are compared with the actual trend of the active cases as shown in Figure 24.

**Figure 24.**
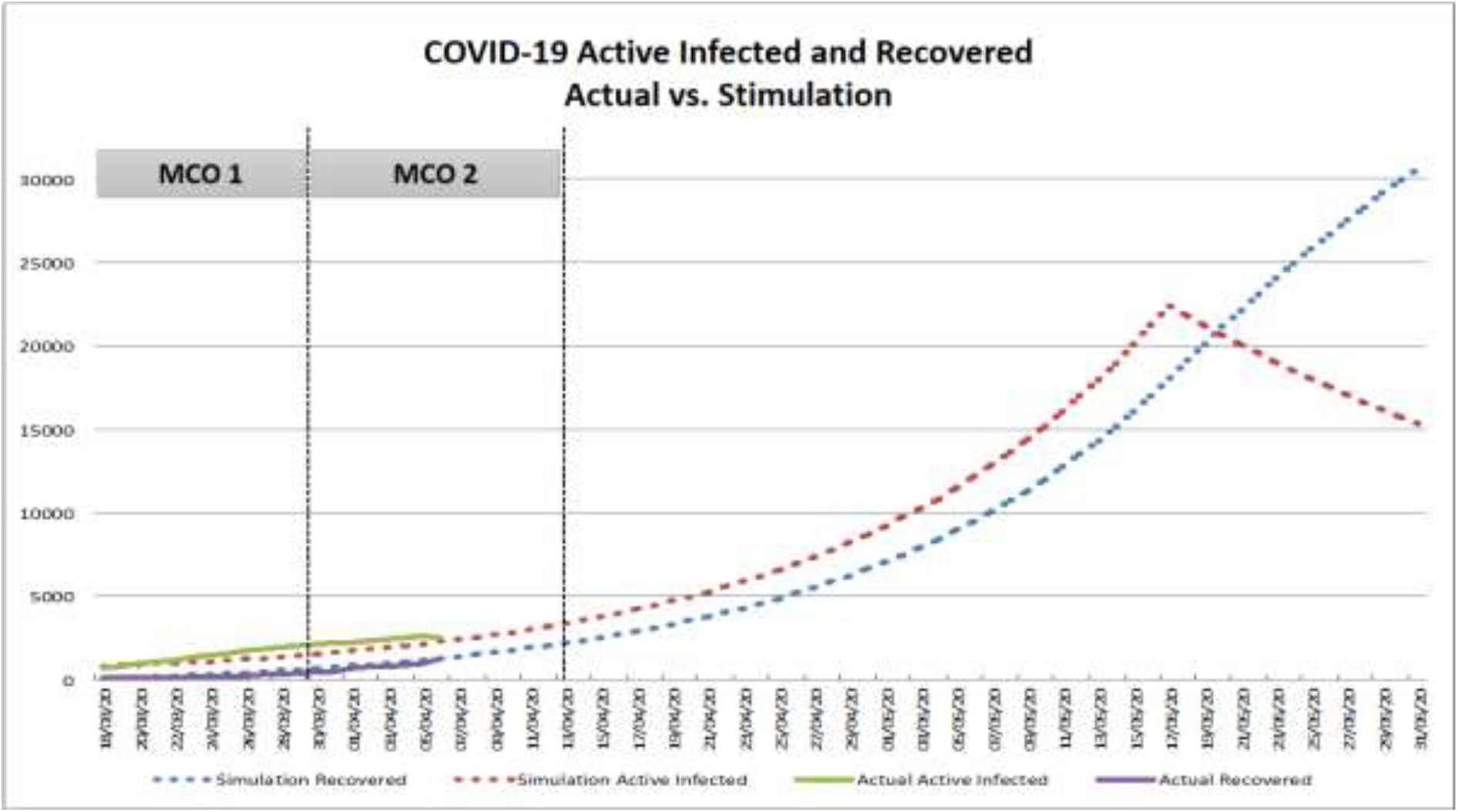
Actual vs. simulated output. Active cases are those still infected (actual infected – recovered-death) with contact rate 1:5 until end of June

Based on Figure 24, the actual active infected and actual recovered are following the projection of the simulation where 1 infected person is still in contact with 5 persons up to end of June before the contact rate is totally restricted to zero. If the actual infected cases and number of recovered continue to follow the projection, there will be a peak of 22,421 active cases on the 17^th^ of May 2020.

Based on this model, our major findings can be summarized as:

1. MCO1 has taken no effect in restricting the contact of an infected person
2. Actual total infected cases are higher than the projection with MCO. Simulation output for total infected with further restriction of contact rate of an infected person (1:1) during MCO2 is lower than actual data (see Figure 19).
3. Actual active infected cases are following the projection of contact rate 1:5 as in Figure 24 (showing low compliance of MCO) which will result in the peak of active infected cases = 22,421 on 17^th^ May 2020.
4. All measures must be taken to make sure that MCO2 compliance is at 100% to avoid higher increase of active cases and to stop the chain of the infection.
5. If the trend of actual data follows the projection in Figure 24, this shows that MCO2 has low compliance. MCO3 must be considered with full enforcement to restrict all the contact to the infected person.

## 4.0 Discussion

Based on the results, we can see that different models have slightly different prediction on the estimated COVID-19 peak dates and a sizeable variation in terms of the maximum number of people infected in Malaysia, as shown in Table 6. Method 1 based on curve-fitting with probability density function estimated that the peak will be reached on 19^th^ April 2020 with an estimation of 5,637 infected persons. Method 2 based on SIR estimated between 630,000 to 800,000 persons will be infected at the peak of outbreak in between 20^th^ to 31^st^ May 2020 if MCO is in place. Method 3 based on Systems Dynamics estimated that 22,421 persons will be infected at the peak date around 17^th^ May 2020. The Mean Absolute Percentage Error (MAPE) was calculated using Eq. 4 and the accuracy reported in the table was calculated using Eq. 5 as reported in Section 3.4 above, based on actual data from 18^th^ March 2020 to 6^th^ April 2020.

**Table 6.**
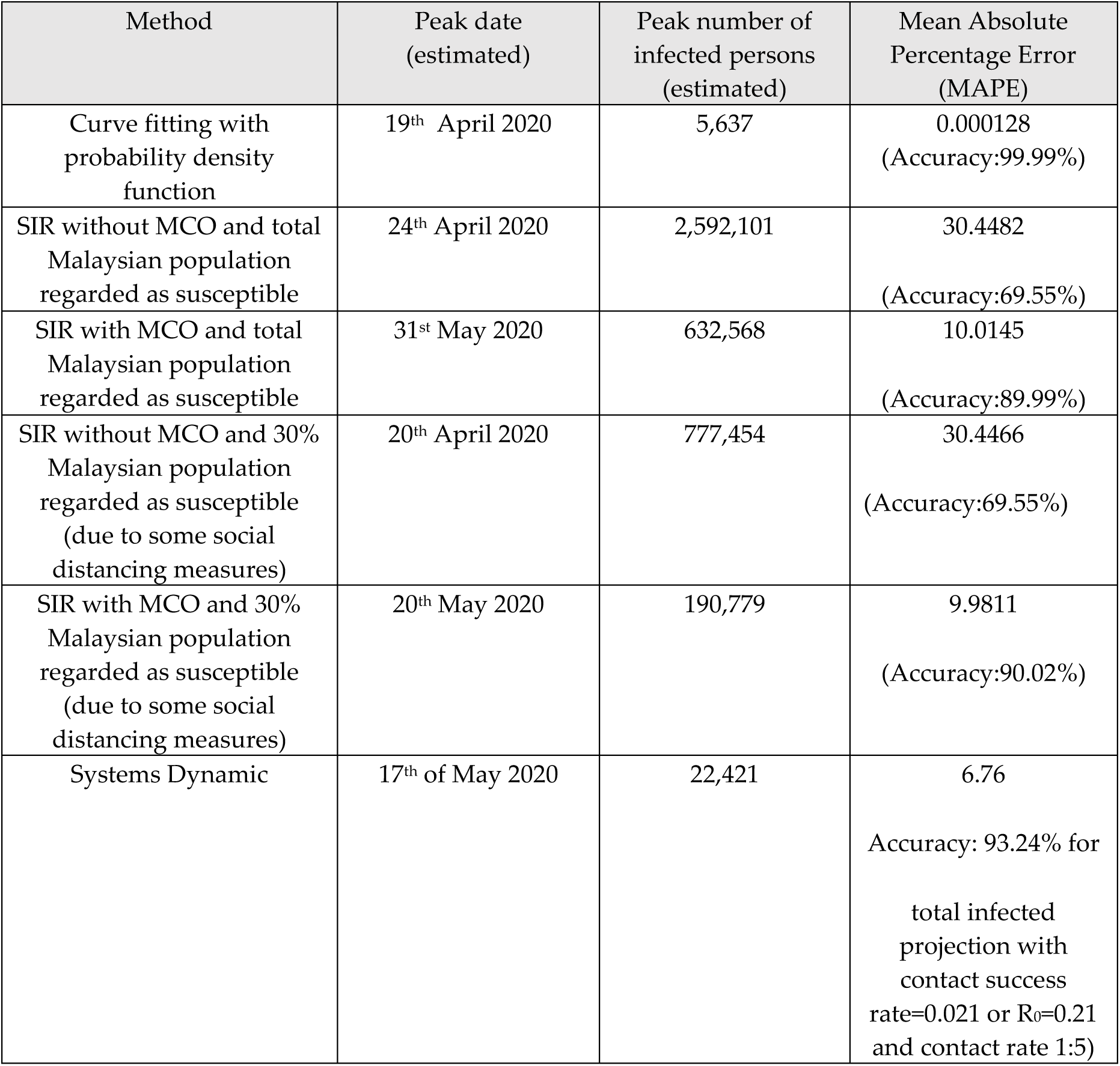
Prediction on the estimated COVID-19 peak dates, maximum number of people infected in Malaysia and Mean Absolute Percentage Error.

| Method | Peak date (estimated) | Peak number of infected persons (estimated) | Mean Absolute Percentage Error (MAPE) |
| --- | --- | --- | --- |
| Curve fitting with probability density function | 19<sup>th</sup> April 2020 | 5,637 | 0.000128<br>(Accuracy:99.99%) |
| SIR without MCO and total Malaysian population regarded as susceptible | 24<sup>th</sup> April 2020 | 2,592,101 | 30.4482<br>(Accuracy:69.55%) |
| SIR with MCO and total Malaysian population regarded as susceptible | 31<sup>st</sup> May 2020 | 632,568 | 10.0145<br>(Accuracy:89.99%) |
| SIR without MCO and 30% Malaysian population regarded as susceptible (due to some social distancing measures) | 20<sup>th</sup> April 2020 | 777,454 | 30.4466<br>(Accuracy:69.55%) |
| SIR with MCO and 30% Malaysian population regarded as susceptible (due to some social distancing measures) | 20<sup>th</sup> May 2020 | 190,779 | 9.9811<br>(Accuracy:90.02%) |
| Systems Dynamic | 17<sup>th</sup> of May 2020 | 22,421 | 6.76<br>Accuracy: 93.24% for<br>total infected projection with<br>contact success<br>rate=0.021 or $R_0=0.21$<br>and contact rate 1:5) |

The models show different modeling have different estimation and vary in terms of difference from actual data produced by Malaysia Ministry of Health. Models to project the progress of COVID- 19 in Malaysia is only as good as the data they are based upon. First, the data needs to be cleaned up so that it reflects the actual numbers. However, epidemiologists have expressed concerns that several factors are affecting the quality of the data. For instance, the number of infected persons per date must be accurate. This data is arguably not accurate when there is a backlog of 8,398 pending COVID-19 test results on April 5, 2020 in Malaysia [7]. As of April 5, a total of 51,937 people was tested, out of which 3,662 tested positive and 39,877 tested negatives. Besides a total of 3,662 confirmed coronavirus cases in Malaysia, the country also reported 61 deaths. Data such as actual dates of when COVID-19 tests are run and dates of when these test results come back is crucial to know the number of people infected on a certain date. Apart from backlog cases that were infected and tested some time ago, it is also uncertain on whether the figures reported each day have excluded data from repeated tests of patients currently under treatment.

In addition, Malaysia are still not doing extensive screening, as tests were only run on individuals that fulfill a specific criterion of potential COVID-19 and contacts of infected patients. It is therefore understandable our estimation might not reflect the actual number of cases that will be published by the Malaysia Ministry of Health in days to come.

Based on the system dynamic model, it is likely that MCO1 has no effect in restricting the contact of an infected person because the actual total infected cases are higher than the projection with MCO. Simulation output for total infected with further restriction of contact rate of an infected person (1:1) during MCO2 is also lower than actual data (see Figure 19). Actual active infected cases are following the projection of contact rate 1:5 as in Figure 24 (showing low compliance of MCO) which will result in the peak of active infected cases = 17,114 on 17th May 2020. All measures must be taken to make sure that MCO2 is 100% complied to avoid the high increase of active cases and to stop the chain of the infection. If the trend of actual data follows the projection in Figure 24, this shows that MCO2 has low compliance. MCO3 must be considered with full enforcement to restrict all the contact to the infected person.

The presence of a significant portion of asymptomatic COVID-19 cases in the community may explain this the lack of effect from MCO1. Asymptomatic cases are not captured because the testing focused more on symptomatic ones. Thus, asymptomatic person easily spread the virus to their household contact without realizing it. This explain why it seems like significant transmissions are still occurring in the community despite stringent social distancing measures. Further actions are needed to ensure that disease control measures during MCO2 will be more extensive.

Relying on stringent social distancing method alone, like MCO, will not be adequate in controlling the outbreak. Basic measures in infectious disease control must be strengthened as these measures play the most important role in this epidemic. Infected person, either asymptomatic or symptomatic must be identified as soon as possible so that the person can be isolated from other susceptible persons. Extensive and rapid contact tracing is needed to identify all exposed person and put under quarantine to stop them from infecting other susceptible persons. Proper surveillance method is needed to ensure compliance. If all these measures are not strengthened, MCO alone will not be sufficient even though it is executed in full force. Nevertheless, MCO is important to reduce massive contacts between peoples so that the healthcare system gained enough time to prepare. It is when all the basic control measures for COVID-19 is strongly grounded and functioning, reflected by down-turn epidemic curve, that’s the moment we can safely lift up the MCO without the fear of recurring big waves of COVID-19 epidemic.

## 5.0 CONCLUSION AND FUTURE WORK

When COVID-19 outbreak was declared as pandemic by WHO, necessary precaution is needed to control the outbreak and addresses the issues that arise. Movement or restricted control order is crucial to mitigate the total number of people infected by COVID-19 and to ensure our health facilities can cope with the number at any given time. In this paper we generate forecasts using a Curve Fitting Model with Probability Density Function and Skewness Effect, the SIR model, and a system dynamic model to predict the trends of COVID-19 cases in Malaysia. We estimated that the peak in terms of the number of infected people will occur between middle of April 2020 to end of May 2020, with total infected numbers varying between 5,000 persons (based on existing number of infected persons) to 2.6 million (based on the total population of Malaysia regarded as people susceptible to the infection). The varying numbers are due to assumptions we have made based on available data.

Compared to actual total infected data from 18/03/20 to 06/04/20, Curve Fitting with Probability Density Function and Skewness Effect Modelling is the most accurate (99.99% accuracy). However, different models used different data, and they may not be accurate due to limited data but they can be used to provide more understanding of the pattern of spread. For instance, the SIR model basically give us an idea where based on the current data, how the outbreak would progress. We tried to measure for both periods before and after the implementation of the MCO. The peak result from the SIR model may not be absolutely accurate due to the limited data. However, the comparison of the modelled progression shows that implementation of the MCO where close contact has been minimized due to the social distancing can flatten the curve. If MCO 3 to be imposed, all measures must be taken to restrict the contact of an infected person as well as their contacts especially those who are under home surveillance and self-quarantine. As shown in the System Dynamic Model, the effectiveness of MCO2 is still not optimum.

Each model has some assumptions, some very much depends on the public’s compliance to the regulations set by the government. The model based on Systems Dynamics can incorporate more parameters in its simulation such as contact rate and duration of recovery to help understand the pattern of spread. The curve-fitting model, despite its reported accuracy, is too dependent on the historical data. The true reflection of reality can only be achieved if more tests can be performed. The SIR model is simple and only considers 3 parameters, which are all inferred from the historical data.

We are now running a prediction model based on deep learning, using data augmentation from other countries. We also plan to combine the models to give a better estimation. A dashboard with adjustable parameters is also currently being developed to interactively understand the effects of different control measures on the projection of infected cases. In the future, we suggest that study be made to decide on the best dates to safely stop our current MCO, to predict the cases according to different time and location and whether different decisions should be made for different states in Malaysia. However, reliable data is needed to enable all these possibilities to be incorporated in our future models and dashboard.

Accurate prediction on the rising and declining period of COVID-19 cases could support the decision for MCO period and the expected level of compliance during the MCO periods. By analyzing the patterns and trends of the cases, forthcoming initiatives can be proposed and implemented.

## Data Availability

I have followed all appropriate research reporting guidelines and other pertinent material as supplementary files, if applicable.

## Acknowledgments

This work is funded by Universiti Teknologi Malaysia under grant no. R.J130000.7751.4J454.

